# Gene-based Hardy–Weinberg equilibrium test using genotype count data identifies novel cancer-related genes

**DOI:** 10.1101/2024.03.20.24304529

**Authors:** Jo Nishino, Fuyuki Miya, Mamoru Kato

## Abstract

**Background:** An alternative approach to investigate associations between genetic variants and disease is to examine deviations from the Hardy–Weinberg equilibrium (HWE) in genotype frequencies within a case population, instead of case-control association analysis. The HWE analysis distinctively requires disease cases without the need for controls and demonstrates a notable ability in mapping recessive variants. Allelic heterogeneity is a common phenomenon in diseases. While gene-based case-control association analysis successfully incorporates this heterogeneity, there are no such approaches for HWE analysis. Therefore, we proposed a gene-based HWE test (gene-HWT) by aggregating single-nucleotide polymorphism (SNP)-level HWE test statistics in a gene to address allelic heterogeneity.

**Results:** This method used only genotype count data and publicly available linkage disequilibrium information and has a very low computational cost. Extensive simulations demonstrated that gene-HWT effectively controls the type I error at a low significance level and outperforms SNP-level HWE test in power when there are multiple causal variants within a gene. Using gene-HWT, we analyzed genotype count data from genome-wide association study for six types of cancers in Japanese individuals and found that most of the genes detected are associated with cancers. In addition, we identified novel genes (*AGBL3* and *PSORS1C1*), novel variants in *CTSO* known to be associated with breast cancer prognosis and drug sensitivity, and novel genes as germline factors, which have associations in gene expression or methylation status with cancers in the combined analysis of six types of cancers.

**Conclusions:** These findings indicate the potential of gene-HWT to elucidate the genetic basis of complex diseases, including cancer.

## BACKGROUND

Case-control association analyses for individual single-nucleotide polymorphisms (SNPs; i.e., single-SNP case-control analysis), such as the chi-squared or Fisher’s exact test on a 2 x 2 contingency table or logistic regression analysis, have been used to assess the genetic association between SNPs and disease states, leading to the detection of numerous disease-related SNPs (1). This approach has been successfully extended to “gene-based” analysis (2-8). Gene-based analysis has several advantages over single-SNP analysis. First, collectively considering multiple variants within a gene may increase the statistical power of the analysis if allelic heterogeneity is present (i.e., different variants at the same gene lead to the same or similar phenotypes). Second, focusing on genes instead of millions of SNPs reduces the burden of multiple tests, which may also increase the power. Third, gene-based analysis addresses the allelic heterogeneity and allows for more consistent findings across different studies on similar diseases. Furthermore, studying genes, the functional units of the genome, can provide valuable insights into the underlying biology of a disease.

Instead of using a case-control analysis, using deviations in genotype frequencies from the Hardy–Weinberg equilibrium (HWE) within a case population, i.e., HWE analysis, is an alternative approach to investigate the association between SNPs and disease (9-12). For a particular locus with two alleles *A* and *a* with frequencies (1 − *p* )and *p*, respectively, the HWE states that the genotype frequencies of *AA, Aa, aa* are (1− *p* )^2^, 2*p* (1 − *p* ), and *p*^2^, respectively, under conditions such as random mating, a large population, and no migration, mutation, or selection (13). Because different genotypes in a disease-causing variant have different levels of susceptibility to the disease, the genotype frequencies within a case population may deviate from the expectations under the HWE, i.e., in Hardy–Weinberg disequilibrium (HWD). Therefore, HWD can be used for genetic association, and this approach, unlike case-control studies, requires only cases and not controls. This method has been used for fine mapping of recessive variants and for providing additional evidence for case-control analysis of recessive variants (9, 12, 14-17).

Analogous to the gene-based case-control analysis, gene-based HWE analysis should be considered owing to its many advantages, including increasing statistical power and the interpretability of results, similar to the gene-based case-control analysis. Multiple recessive mutations commonly exist within the same disease-causing gene, (18) and the gene-based HWE analysis is precisely targeted for such scenarios. However, until now, no such method has been proposed.

Therefore, we proposed a gene-based HWE test (gene-HWT), which advantageously uses genotype count data and publicly available linkage disequilibrium information, without requiring individual genotypes, from genome-wide association studies (GWAS) with increasingly large sample sizes (19). Note that this proposed use of HWD is not intended to identify genotype errors as is commonly done (20). Rather, the use of data in which mutations with a high probability of error have been removed prior to analysis is intended. The results obtained did not show any features attributable to errors.

## RESULTS

### gene-HWT

We started with the statistic for the single-SNP Hardy–Weinberg equilibrium test (single-SNP HWT) and then introduced the proposed method, gene-HWT (an overview is illustrated in Fig. 1). For a particular locus with two alleles *A* and *a* with frequencies *q*= 1 − *p* and *p*, respectively, the statistic for single-SNP HWT, *z*, in *n* diploid samples is calculated as follows:

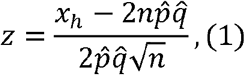

where *x*_*h*_ is observed number of heterozygosity in the sample, and 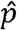 and 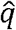 represent the sample frequency of *a* and *A*, respectively (11). Under HWE, *z* is expected to be 0 since the genotype frequency of *Aa* is expected to be 2*npq*. HWT is performed based on the fact that *z* asymptotically follows a standard normal distribution under HWE. Note that the commonly used statistics for single-SNP HWT is the square of *z, z*^2^ (11).

**Figure 1.**
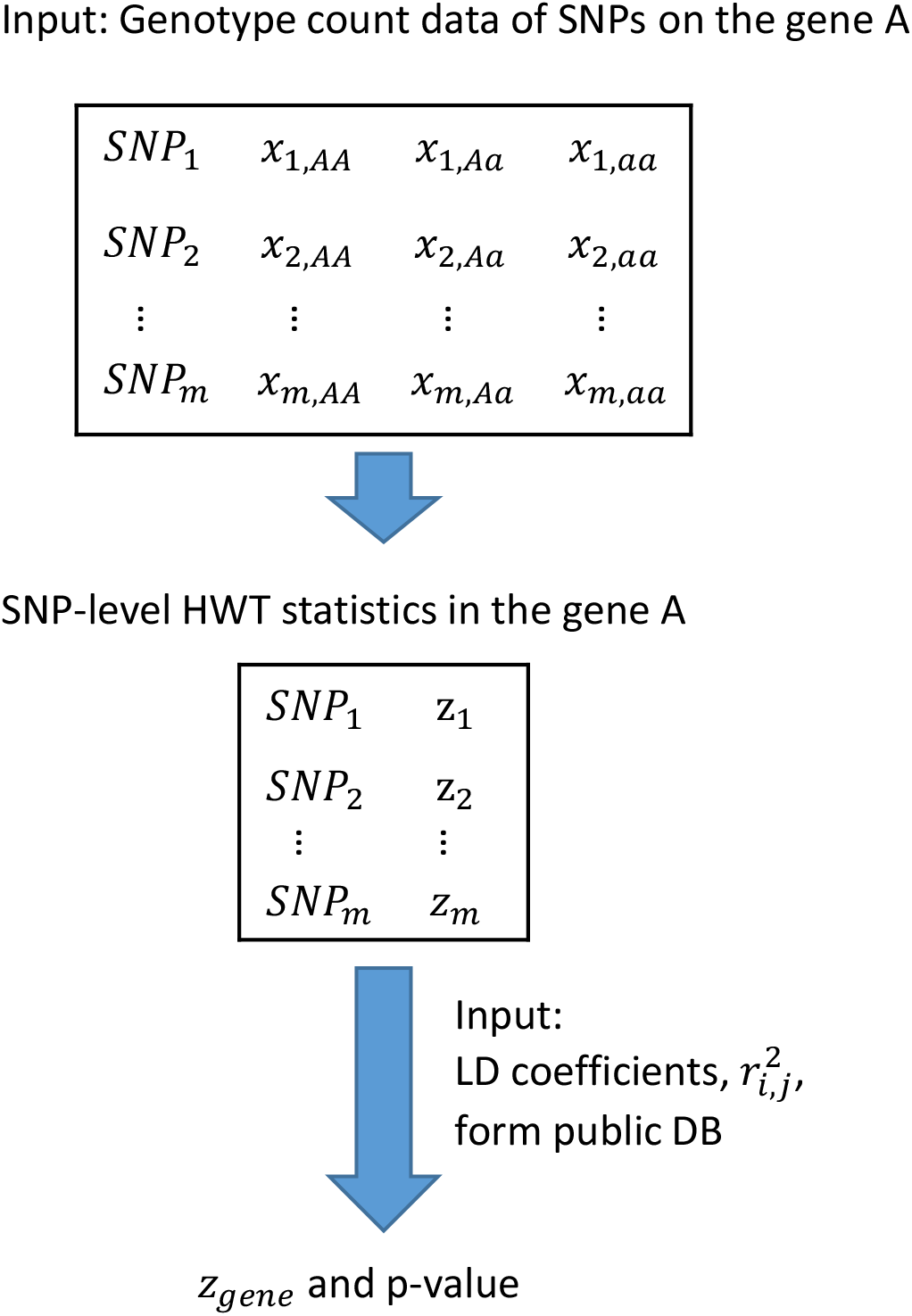
Overview of the gene-HWT. The input is genotype count data of SNPs located on a specific gene. Within this gene, SNP-level HWT statistics are computed. To consider the correlations among *z*′_*i*_*s*, linkage disequilibrium (LD) coefficients, 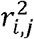, are derived from a public database. The gene-based HWT statistics, *z*_*gene*_, and corresponding p-value are calculated.

The test using *z* employs continuous approximation, which does not yield appropriate results when the number of minor alleles in the sample is low. Therefore, in this study, we focused on loci with a minor allele frequency (MAF) ≥5% in the sample. In addition, Yates’ continuity correction was applied to *z* when the expected number of homozygotes for minor allele was ≤5. The correction was performed by subtracting 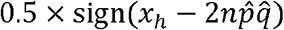 from the numerator of *z*, where sign() returns the sign of a real value.

For a gene with *m* loci, we proposed the statistic for gene-HWT, *z*_*gene*_, as

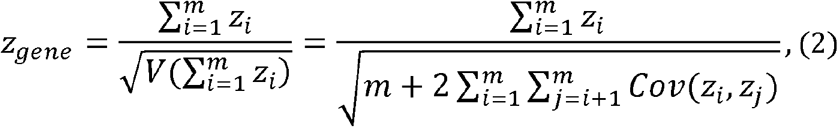

where *z*_*i*_ is the HWT formula for *i*-th variant in the gene *z*_*gene*_ is the sum of *z*_*i*_ divided by its standard deviation (Fig. 1), enhancing the detection of cumulative accumulation of homozygote or heterozygote excesses within a gene. This statistic includes the covariance between *z*_*i*_ and *z*_*j*_, *Cov* (*z*_*i*_, *z*_*j*_), due to LD, making the direct computation of *z*_*gene*_ challenging. Considering the representation of *Cov*(*z*_*i*_, *z*_*j*_) in terms of LD coefficients *r*_*i,j*_, between the *i*-th and *j*-th variants, we successfully proved 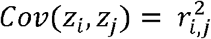 as *n* is large in a Supplementary Note. Therefore, *z*_*gene*_ is as follows:

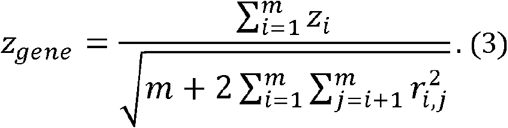

The 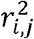 values were retrieved from a public database. Therefore, to calculate *z*_*gene*_, only the genotype counts were required. The gene-HWT was performed using the standard normal distribution: since *z*_*gene*_ is the standardized sum of normal variables, *z*_*i*_, *z*_*gene*_ asymptotically follows a standard normal distribution under the null hypothesis that all *m* variants in the gene are under HWE.

### p-values under the null model and type I error rates

Under the null hypothesis (HWE), the behavior of the p-value and the type I error rates of gene-HWT were investigated by simulation. In each simulation, one gene was randomly selected, with replacement, from 388 genes that meet certain criteria on chromosome 20 from the 1000 Genomes Phase 3 (21) dataset (see Methods for details). Using Hapsim (22), *n* diplotypes are generated while preserving the real LD structure. The QQ-plots displayed p-values obtained from 20,000 simulations for each setting, representing approximately the total number of genes in the human genome (Fig. 2). The observed – log_10_(P) values obtained from gene-HWT (Fig. 2, circles) exhibited a good fit to the theoretical straight line under HWE for all sample sizes, *n* = 200, 1000, and 3000. In contrast, when LD was not corrected, i.e., when using the statistic with LD set to 0 in (2), the observed – log10(P) values (Fig. 2, cross) were substantially inflated from the expected theoretical curve, leading to the inflation of type I error rates.

**Figure 2.**
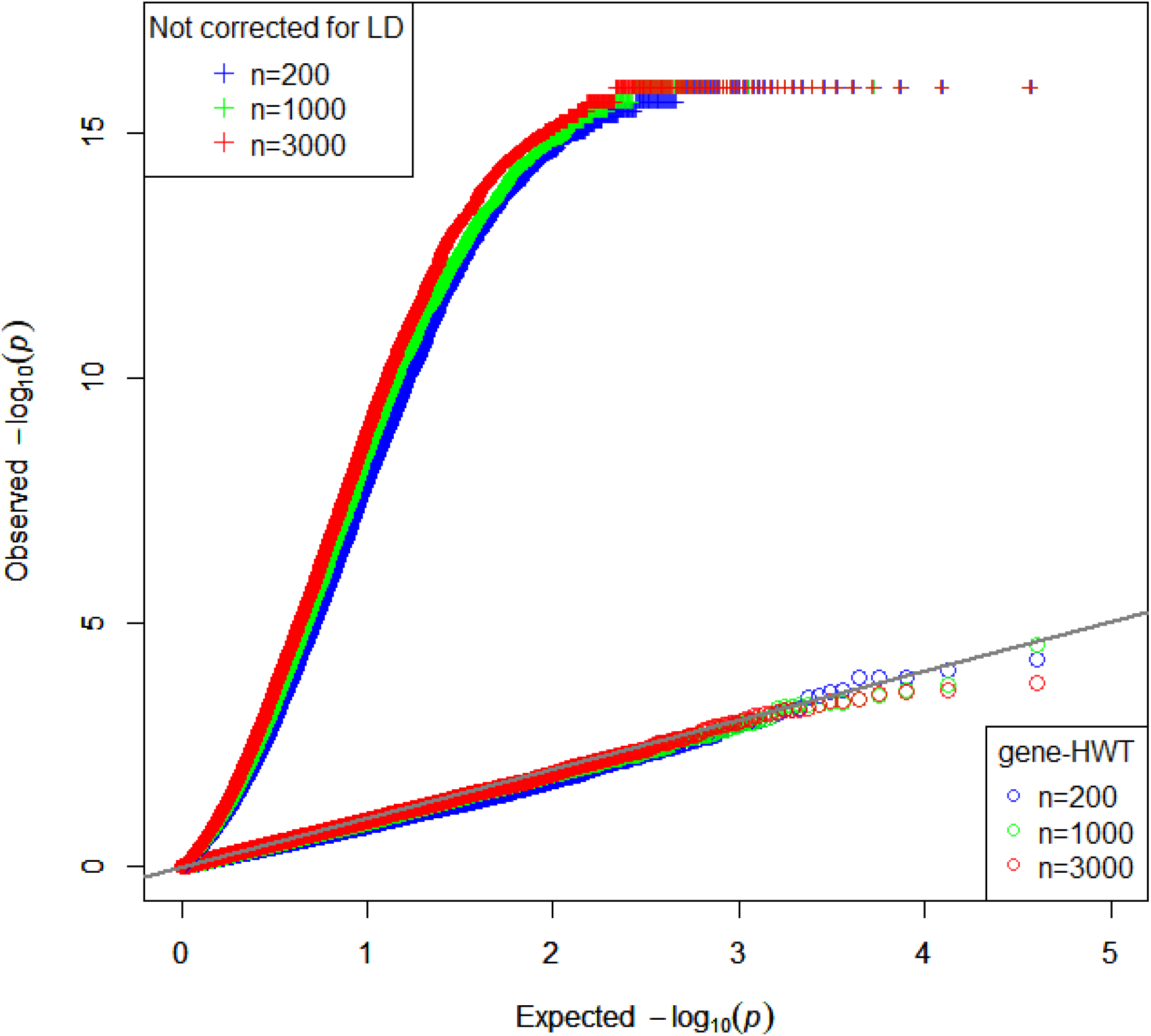
QQ plot of P-values for gene-HWT and the test not corrected for LD under the null hypothesis. P values for gene-HWT and the test not corrected for LD through simulations with sample size (n)=200, 1000 and 3000 under the null hypothesis (HWE), utilizing real data from chromosome 20 of the EAS population in the 1000 Genomes Phase 3. The grey line represents the expected value under the null hypothesis.

Type I error rates by one million simulations for each setting are presented in Table 1. When LD was not corrected, the type I error rate was much larger than the nominal significance level. Type I error for gene-HWT tended to be conservative when the sample size was small, especially when the nominal significance level was large. For example, the type I error rate was 3.0% under *n* = 200 and α = 5%. When the sample size was large, *n* = 1000 or 3000, especially with a small nominal significance level, the type I error rates of gene-HWT were very close to the nominal significance level. At the nominal significance level of 0.025%, corresponding to Bonferroni-corrected 5% significance level for 20,000 genes in the human genome, the type I error rates were 0.022% and 0.026% under *n* = 1000 and *n* = 3000, respectively. Therefore, even at small significance levels, such as those used in the genome scan, the gene-HWT type I error rate can be effectively controlled by appropriately adjusting the LD.

**Table 1.**
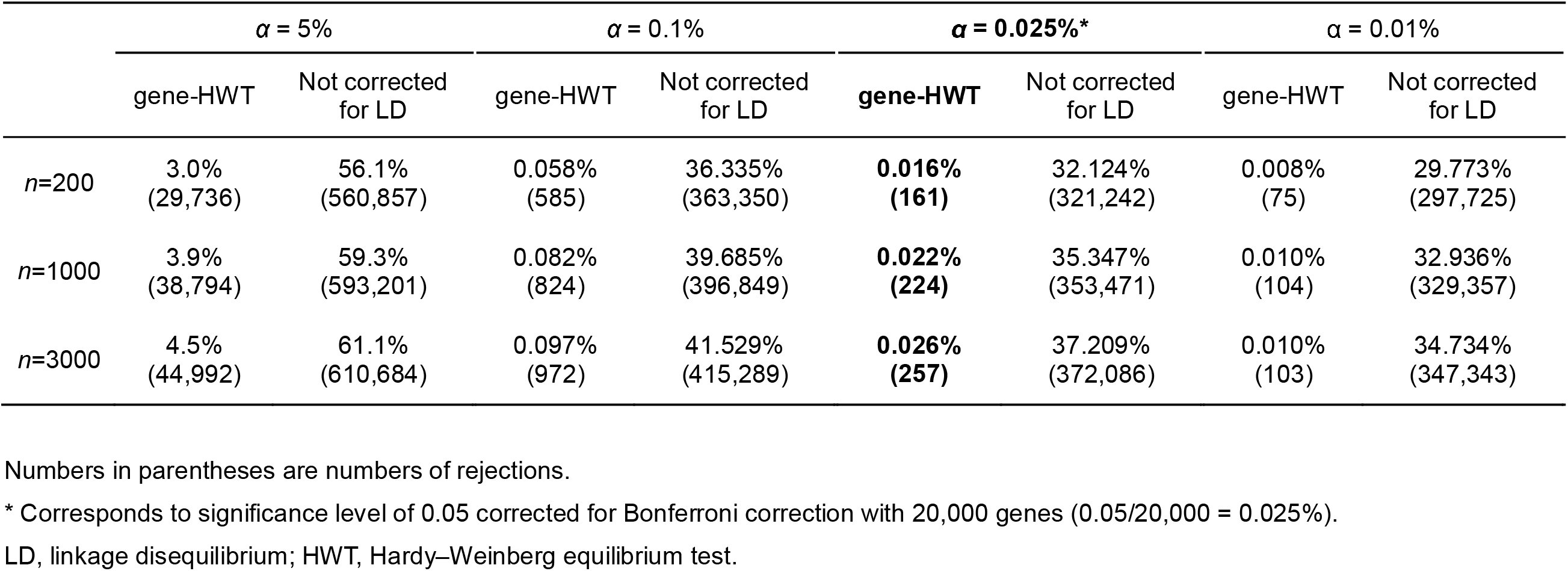
Empirical type I error rates from 1,000,000 simulations.

### Power

A power analysis was conducted for gene-HWT under a multiplicative relative risk model, with 1-12 causative SNPs randomly assigned within a single gene. Diplotypes for genes on chromosome 20 were simulated in the same way as examining type I error rates. The genotype risk ratios for a causal SNP was defined as *AA* : *Aa* : *aa* = 1 : (1+*β*_1_) : (1+*β*_2_). The individual’s relative risk was obtained by multiplying the risk ratios of each variant. The absolute risk was proportional to the relative risks, under the constraint of a prevalence (average risk) of 0.1 (see Methods for details).

The powers for gene-HWT are shown in Fig. 3. The recessive (2*β*_1_ = 0, *β*_2_ > 0) and dominant (2*β*_1_ *= β*_2_ > 0) models were as follows. A larger sample size increased the power. More causal SNPs led to greater detection of power. Even a small increase from 1 to 3 causal SNPs significantly increased the power of detection. For example, when *β*_2_ = 0.2 in a recessive case, with *n* = 200, 1000, and 3000, the detection rates increased from 2.8% to 13% (4.64-fold), 8.7% to 36.9% (4.24-fold), and 18.5% to 69.4% (3.75-fold), respectively. In both recessive and dominant models with sufficient sample size (*n =* 3000), even for relatively weak effects with *β*_2_ = 0.05, 0.1 and 0.2, a power of 70% was achieved with 12, 6, and 3 causal SNPs, respectively. Under the same value of *β*_2_, the power for the recessive and dominant models were equivalent but deviated in opposite directions from HWE, with the recessive model showing “Homozygote excess” (z < 0) and the dominant model showing “Heterozygote excess” (z > 0) (Fig. 4). The semidominant model (2*β*_1_ *= β*_2_ > 0) had very low power.

**Figure 3.**
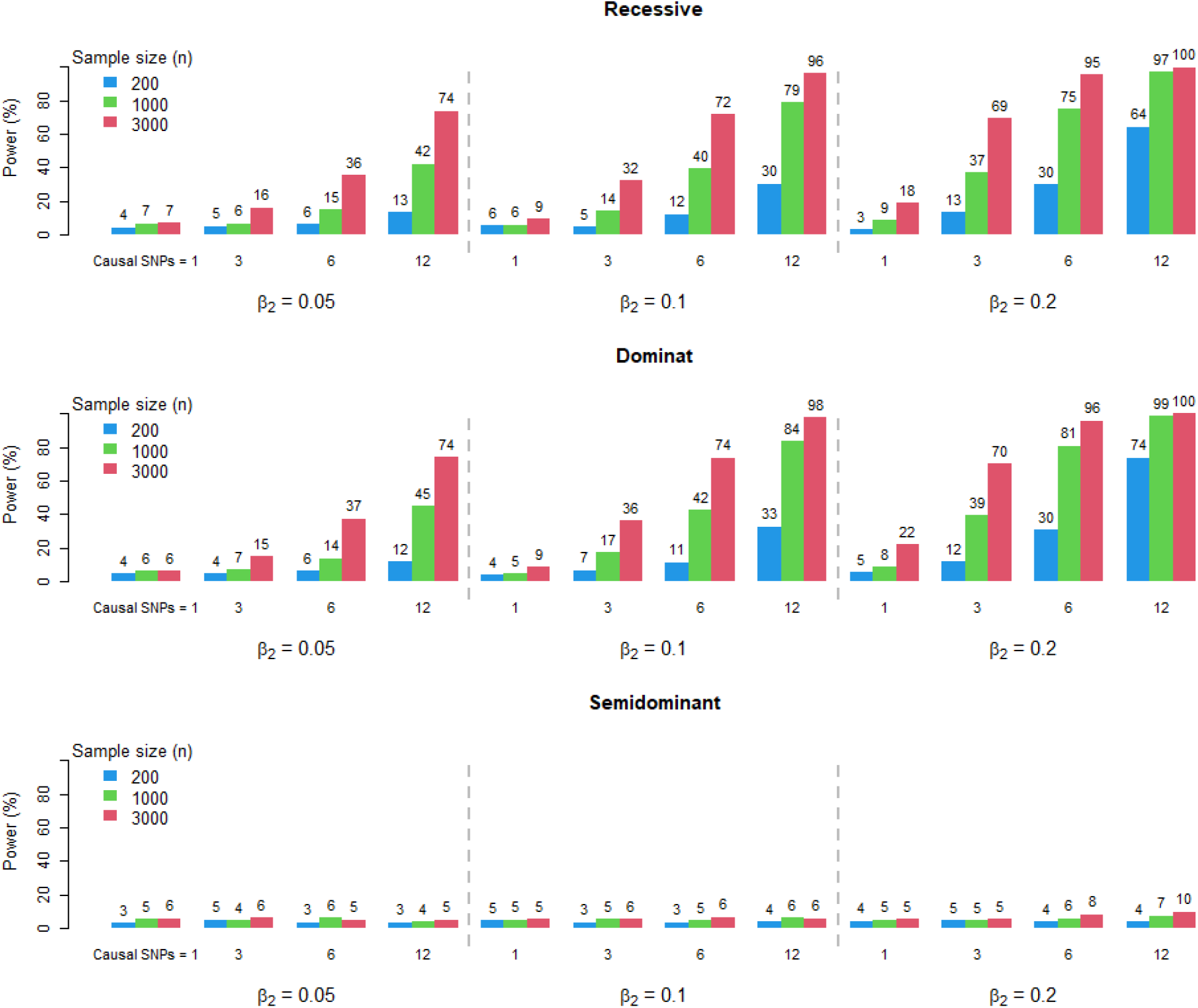
Power of gene-HWT. The results shown are based on simulations with sample size (n)=200, 1000 and 3000 using data from chromosome 20 of EAS population in the 1000 Genomes Phase 3. The genotype risk ratios for a specific causal SNP are defined as AA : Aa : aa = 1 : (1+*β*_1_) : (1+*β*_2_). Simulations were performed for the recessive model (*β*_1_=0, *β*_2_>0), dominant model (*β*_1_=*β*_2_>0), and semidominant model (2*β*_1_=*β*_2_>0).

**Figure 4.**
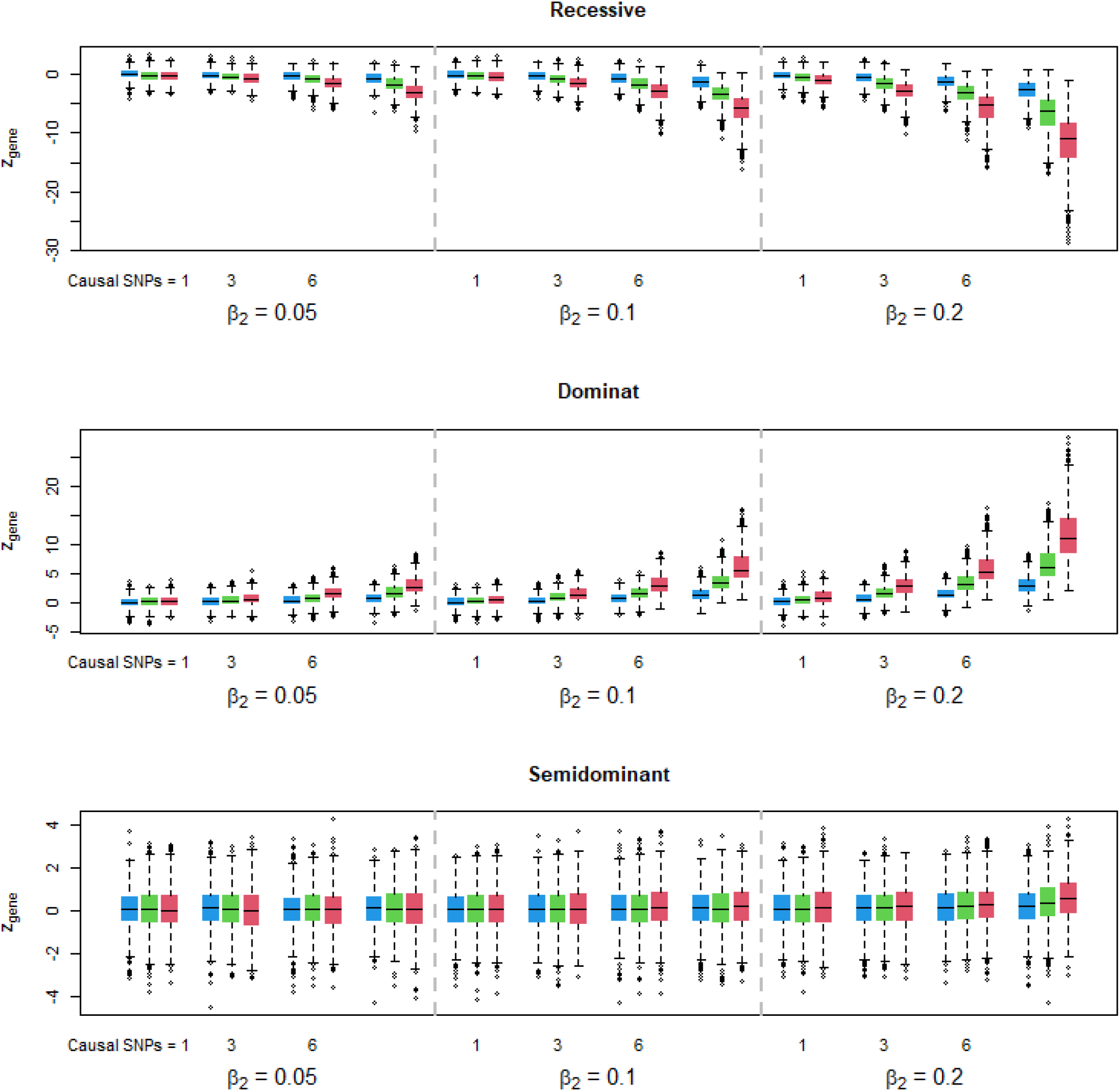
z-values of gene-HWT (z_gene_). z-values of GHWT obtained from the same simulations as in Figure 2.

### Power comparison: gene-HWT versus single-SNP HWT

A comparison of the power of gene-HWT and single-SNP HWT, at the overall significant level of 0.05 both with multiple testing corrections, is shown in Supplementary Fig. 1. Specifically, for each parameter set, 1000 genes (=1000 simulations) were set and in gene-HWT Bonferroni correction was applied for the testing of 1000 genes. In single-SNP test, Bonferroni correction was applied for the number of SNPs (on average, 78,537 SNPs across parameter sets) within each of the 1000 genes.

Compared with the single-SNP test, gene-HWT generally exhibits higher power (Supplementary Fig. 1). Particularly, in cases of intermediate detection power, gene-HWT exhibits a detection power approximately 1.2 to 1.8 times greater than that of the single-SNP testing. For example, when *n* = 200, causal SNP = 12, and *β*_2_ = 0.2 in the dominant model, the power of single-SNP test was 14.4%, while that of gene-HWT was 25.6% (1.78-fold increase). In the recessive model, the power of single-SNP test was 15.9%, whereas that of gene-HWT was 22.8% (1.43-fold increase).

The power of detection was compared between single-SNP test and gene-HWT using the standard genome-wide significance levels as shown in Supplementary Fig. 2. In gene-HWT, the number of genes was set to 20,000, corresponding to Bonferroni-corrected significance level of P < 2.5 × 10^-6^. For single-SNP test, we assumed 1 million SNPs, corresponding to Bonferroni-corrected significance level of P < 5 × 10^-8^. The results aligned with the previous comparison (Supplementary Fig. 1), demonstrating that gene-HWT generally displays a higher power of detection than single-SNP test.

### Analysis of genome-wide data in six cancer types

The genotype count data from GWASs for esophageal, lung, breast, gastric, colorectal, and prostate cancers in Japanese individuals were obtained from the website of the National Bioscience Database Center (NBDC) Human Database (23). Each dataset had been quality-controlled and consisted of approximately 190 individuals. LD information was obtained from the 1000 Genomes Phase 3 dataset (21). The SNPs overlapping with genes (within 2 kb upstream or downstream of the transcripts), and those with MAF ≥5% were selected. For the six types of cancers, gene-HWT was applied to analyze 11,813 to 13,482 genes and 92,690 to 174,270 SNPs (see Methods for details).

Eighty cancer type-gene pairs were identified by applying the gene-HWT with the criterion: FDR q-value < 0.2 for each cancer type (Supplementary Table 1). The 80 genes encompassed SNPs ranging from 1 to 103. To identify common causal genes in cancers, the results of six cancer studies were combined. In the combined analysis (see Methods for details), 11 genes were identified with q-values < 0.05 (Table 2). The combined *z*_*gene*_ values are all negative, suggesting that these 11 genes may have recessive mutations. Among these 11 genes, 8 genes (*CCDC32, POFUT2, PPP1CB, QRFPR, FSTL4, ACRV1, CTSO*, and *GPR180*) have been reported to be associated with gene expression, methylation, or germline mutations, and 3 genes, including *AGBL3* and *PSORS1C1*, were newly identified as candidate cancer-related genes. Additionally, among the genes detected using the threshold of FDR q-value < 0.2 for each cancer type (Supplementary Table 1), 8 genes were found in multiple cancers (Table 3). For genes except for *OR4N2, z*_*gene*_ values were negative, suggesting the presence of recessive mutations. *HLA-DMA* was identified in four cancer types, while *ZNF736* and *AL163636*.2 were identified in three cancer types, and the remaining genes were observed in two cancer types.

**Table 2.**
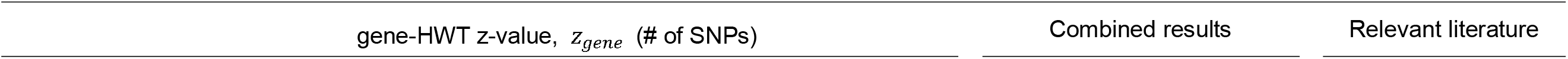
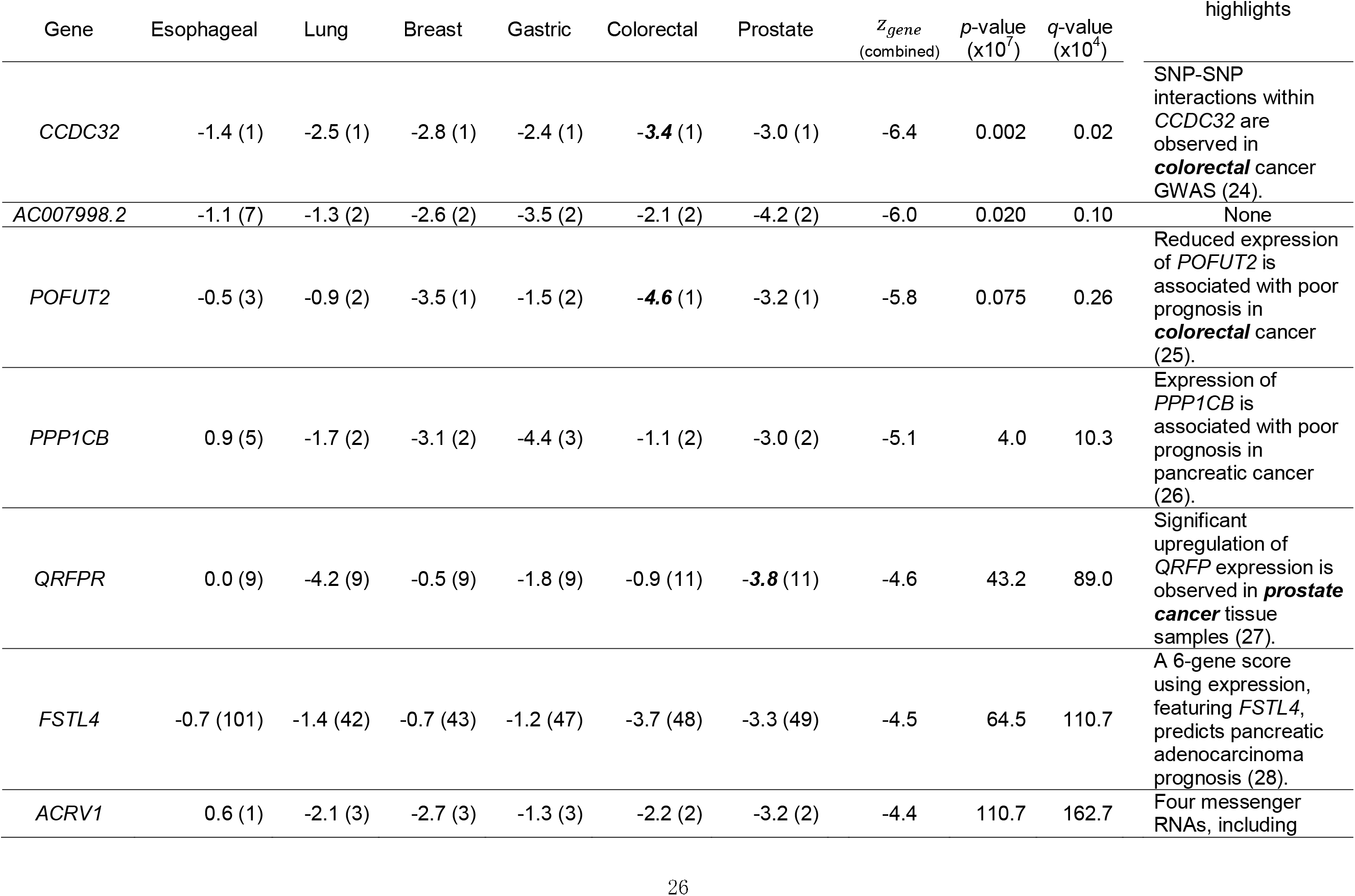
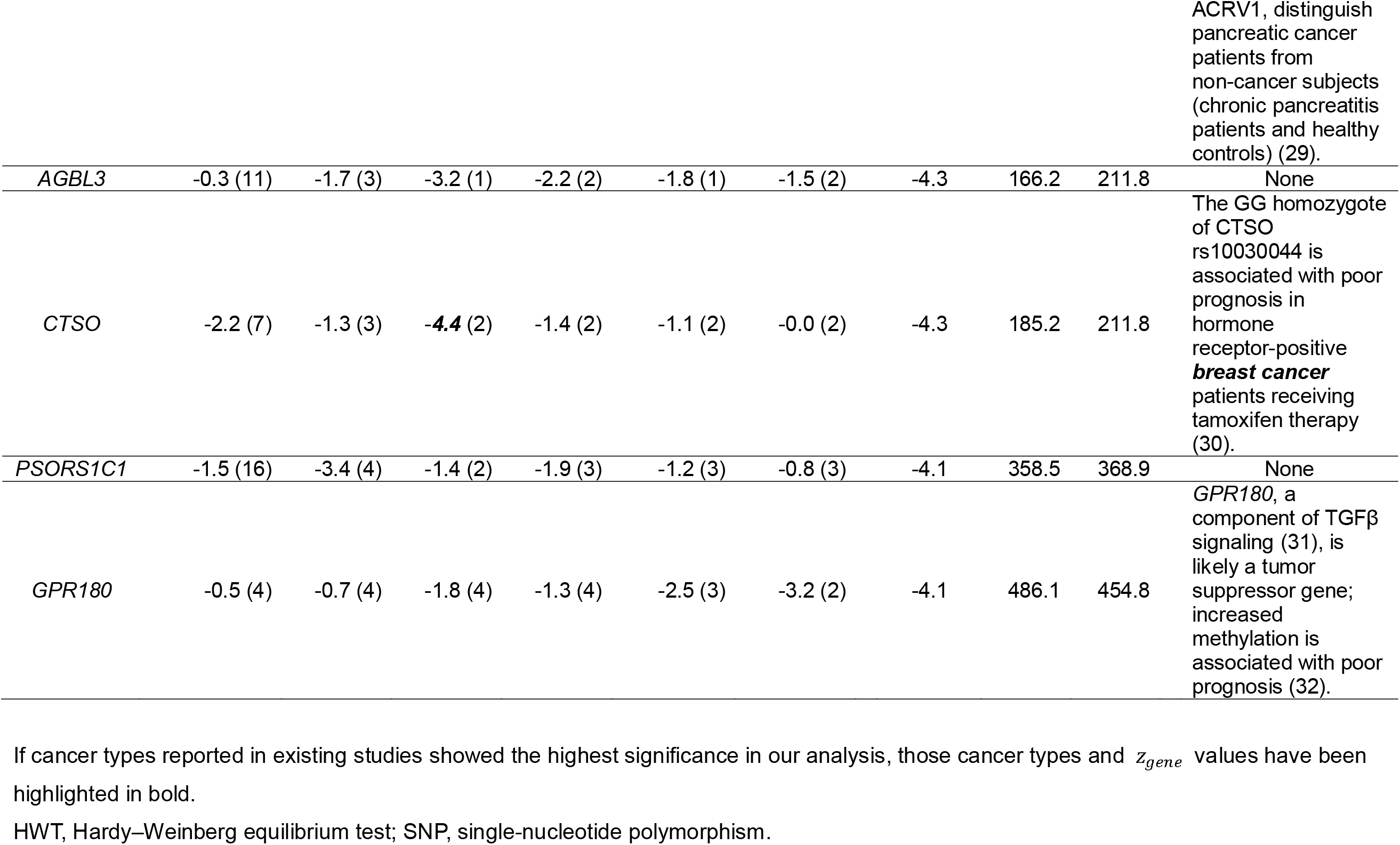
Genes identified by combined gene-HWT results for six cancer types at *q*-value < 0.05.

**Table 3.**
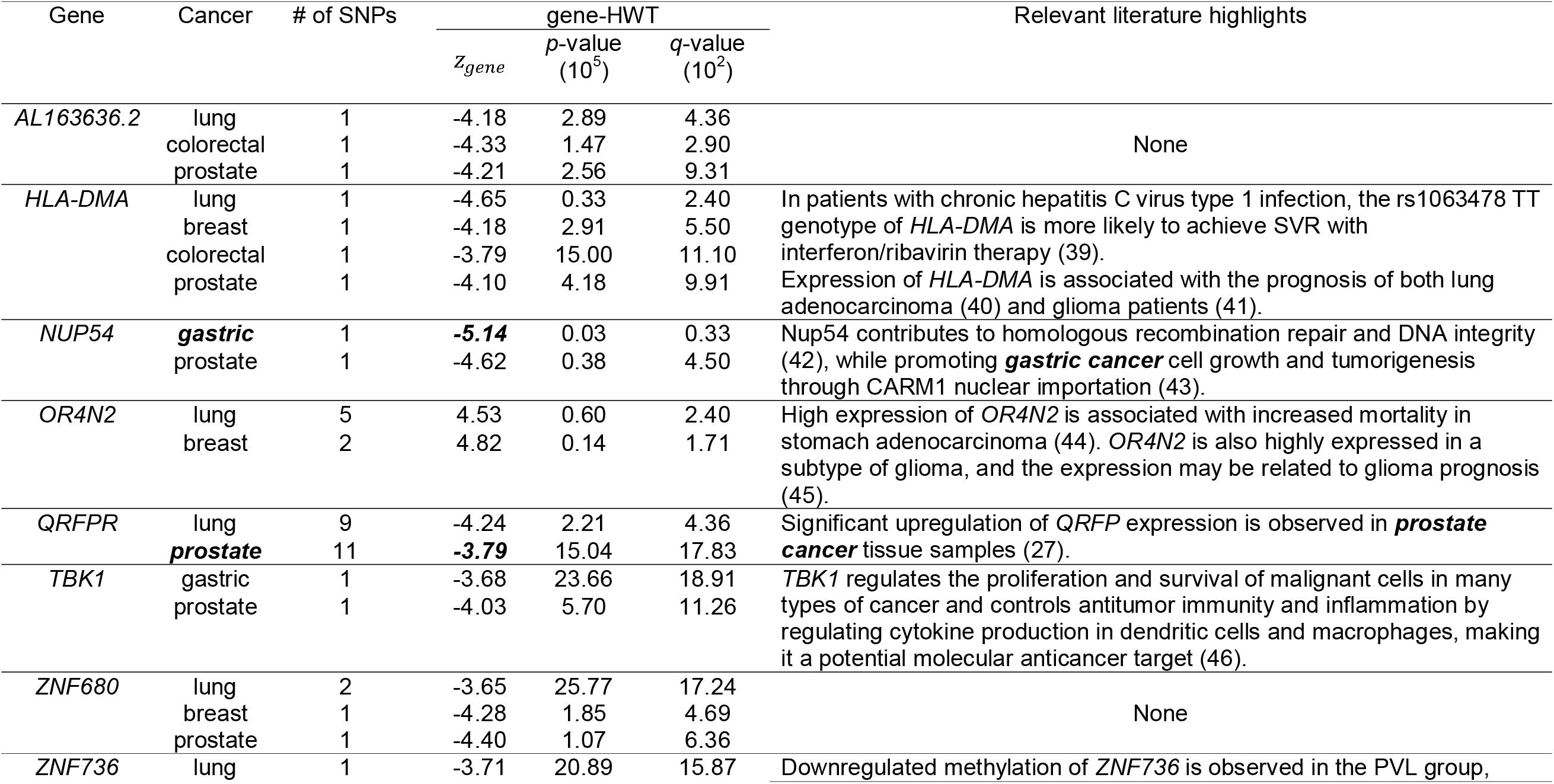
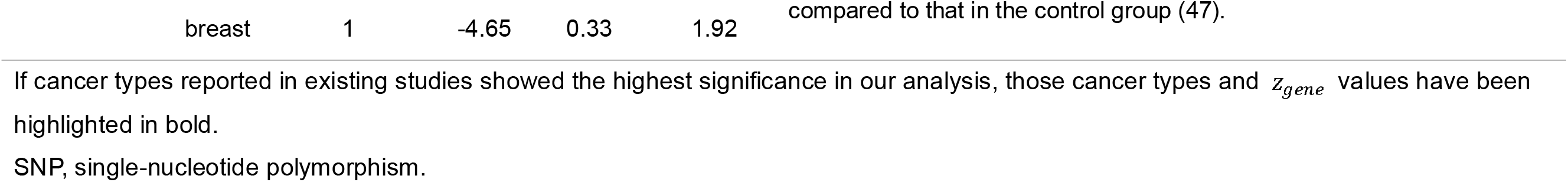
Genes detected in multiple cancer types with FDR q-value < 0.2 criteria.

## DISCUSSION

The proposed method, gene-HWT, is the first method to detect HWD at the gene-level by aggregating HWD in genetic variants (SNPs) within or close to the gene, while adjusting the LD among variants. This test uses only genotype count data, without the individual genotype data, and publicly available LD information. The derived simple relationship between the covariance of the HWT statistic of a pair of variants and the LD coefficient, i.e., 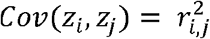, allows for the immediate calculation of the gene-HWT statistic from the single-SNP HWT statistics in a gene of interest without computationally intensive permutation or simulation. gene-HWT effectively controls the type I error rate at a very low significance level, enabling genome scanning, and exhibits significantly increased power compared to single-SNP tests as the number of causal variants within the gene increases. The method also allows for combining results for each gene from different studies, i.e., studies for different cancer types as shown in this study, with allelic heterogeneity, which might lead to further increases in power.

We applied the gene-HWT to six cancer genome data sets, each with approximately 190 cases. By combining these results, we identified 11 significant genes at a threshold of q-value = 0.05. Of the 11 significant genes, 8 genes (*CCDC32, POFUT2, PPP1CB, QRFPR, FSTL4, ACRV1, CTSO*, and *GPR180*) were reported to be associated with various cancers (24-32). Furthermore, of those eight genes, cancer types reported in existing studies often showed the highest significance in our analysis. These findings suggest that gene-HWT captures true causal genes.

*CTSO* exhibited the most significant association with breast cancer (*z*_*gene*_ = –4.44) in a recessive manner in the combined analysis. This aligns with previous reports indicating that the homozygous G variant of *CTSO* rs10030044 is associated with a worse prognosis (i.e., in recessive form) in patients with hormone receptor-positive breast cancer receiving tamoxifen therapy (30). The SNP rs10030044 and the linked rs4256192 lead to increased *CTSO* expression, resulting in decreased *BRCA1* expression and, consequently, tamoxifen resistance (33). Our results showed that *CTSO* could be one of the causative genes for not only breast cancer but also other types of cancer, particularly esophageal cancer, as indicated by its large negative z-score (*z*_*gene*_ = –2.2). For breast cancer, the strong effect was observed with the SNP rs10019161 (single-SNP HWT *P* = 2.18 × 10^-9^), which was also significant in lung cancer (*P* = 0.0054). In esophageal cancer, the strong effects were observed in rs10019975 (*P* = 0.0174) and rs7684248 (*P* = 0.0563). According to the LDmatrix (34) using data in the 1KG Project EAS population (21), these three SNPs rs10019161, rs10019975 and rs7684248 are not in LD with the previously reported rs10030044 and rs4256192 (*r*^2^ < 0.01). This suggests that these three *SNPs* (which are in LD with each other with *r*^2^ > 0.4) could potentially act as independent factors in the cancer of the two previously reported SNPs.

In the analysis of each of the six cancer types in Japanese individuals, the positive z-score for *TP53* detected only in colorectal cancer (*P* = 1.45 x 10^-4^), suggesting dominant variants, could be due to “dominant negative” features for *TP53* variants (35). Furthermore, *RAD51* (36), *APOBEC3A* (37), and *BRAP* (38) identified in gastric, lung, and esophageal cancers, respectively are well-known for their association with cancer. In the analysis of each of the six cancer types with a threshold of q-value = 0.2, out of the 8 detected genes, 6 genes (*HLA-DMA, NUP54, OR4N2, QRFPR, TBK1* and *ZNF736*) are associated with cancers (27, 39-47). *HLA-DMA* showed significant associations with four types of cancer in this study. HLA-DM (encoded by *HLA-DMA* and -*DMB* ) is a non-classical MHC class II-like protein that acts as a peptide editor in the antigen presentation process and plays a crucial role in regulating the loading of antigenic peptides onto MHC class II molecules. In patients with chronic hepatitis C virus type 1 infection in the Chinese Han population who underwent interferon/ribavirin therapy, individuals carrying the rs1063478 TT genotype of *HLA-DMA* had a higher likelihood of achieving sustained virological response (SVR) (39). In patients with lung adenocarcinoma, low *HLA-DMA* expression was associated with disease-specific survival and overall survival (OS) (40). In glioma, high *HLA-DMA* expression was associated with poor prognosis (41). These findings and the result of our analysis suggest that *HLA-DMA* may be associated with a variety of cancers.

New candidate cancer-related genes were identified in our analysis, such as *AGBL3, PSORS1C1*, and *ZNF680*. ATP/GTP Binding Protein-Like 3 (AGBL3), is a metallocarboxypeptidase that mediates deglutamylation of both tubulin and non-tubulin target proteins. Psoriasis Susceptibility 1 Candidate 1 (*PSORS1C1)* is one of the genes associated with psoriasis (48), an inflammatory skin disease, and since there is a known association between psoriasis and cancer (49), PSORS1C1 may also be a cancer-causing gene. In addition, many previous reports have focused on mRNA expression and methylation studies, and this is the first study to find an association between SNPs and cancer for genes such as *OFUT2, PPP1CB, QRFPR, FSTL4, ACRV1, GPR180, OR4N2, QRFPR*, and *ZNF736*.

The public data of the six cancer types used in this study had been quality controlled, and thus, HWD detected by gene-HWT was not attributable to genotyping errors. If the effect of error had been dominant in the detected genes, a tendency towards excess heterozygosity would have been observed (20); however, many detected genes showed excess homozygosity. This is not likely due to population structure or inbreeding (50, 51). If the effect had been large, the z-value should be negative overall, but the median SNP-level z-values for the six cancers were close to zero: –0.021, –0.026, –0.015, –0.056, and –0.029 for esophageal, lung, breast, stomach, colon, and prostate cancers, respectively. Of course, in the case of non-negligible effects of population structure or inbreeding, gene-HWT may produce erroneous results. Thus, the development of a gene-based HWE test that considers population structure and inbreeding is a future challenge.

The proposed method has certain limitations. It targets common variants with MAF ≥5%. As a result, many variants would be excluded from consideration. BRCA1- and BRCA2-associated hereditary breast and ovarian cancer (HBOC) follow a dominant inheritance pattern. Such dominant variants exert their effects heterozygously, making it difficult for them to be highly maintained in the population through natural selection. Indeed, all genes detected, except for *ORN2*, exhibited negative *z*_*gene*_ values, indicating the presence of recessive mutations in those genes. The reason for excluding variants with MAF <5% is that the single-SNP HWT may not work well with rare mutations owing to breakdown of continuous approximation, and naturally, gene-HWT would also fail for these cases. Moreover, since gene-HWT enhances the detection of cumulative accumulation of homozygous or heterozygous excess within a gene, it might be difficult to detect genes with both recessive and dominant mutations using gene-HWT.

## CONCLUSIONS

In summary, we proposed a novel method for detection of gene-based HWD, which uses only the genotype counts and publicly available LD information. It is common for specific genes to have multiple disease-causing mutations, and our approach can aggregate their cumulative effects to enhance the detection power. We successfully demonstrated the application of this method on cancer genomic data, showing its effectiveness. Together, these findings highlight the potential utility of gene-HWT in elucidating the genetic basis of cancers and other complex diseases.

## METHODS

### Simulation for type I error rates of gene-HWT

Type I error rates for the proposed gene-HWT were investigated by simulations under the null hypothesis (HWE) using real data for mimicking realistic LD structure. Specifically, phased genotype data from chromosome 20 in the East Asian (EAS) population, comprising 504 individuals from the 1000 Genomes Phase 3 (21), were utilized. Only SNPs with MAF ≥5% were selected. To reduce the computational burden and to specify a maximum of 12 causal SNPs for subsequent power analysis, genes with 12 to 200 SNPs on chromosome 20 were selected, resulting in the use of 388 genes. In each simulation, one gene was randomly selected from 388 genes obtained, and using Hapsim (22), 2n haplotypes were generated while preserving the LD structure obtained from real data within the gene. Then, 2n haplotypes were randomly combined to create n diplotype and finally, gene-HWT was applied.

### Simulation for power analysis of gene-HWT

A power analysis was conducted using simulations based on a disease causation model involving 1-12 causal SNPs in a single gene. The process of creating diplotypes was identical to that of simulation for type I error rates. The causal SNPs were randomly determined in SNPs within the genes. The genotype risk ratio for each causal mutation is defined as *AA* : *Aa* : *aa* = 1 : (1+*β*_1_) : (1+*β*_2_). The individual’s risk ratio was determined by multiplying the risk ratios for each variant. The individual’s absolute risk was determined while considering the constraint of prevalence = 0.1. In one simulation, a sufficiently large population with ‘*n*/prevalence’ diploids was created in advance, and then *n* individuals were selected based on each individual’s absolute risk. Finally, gene-HWT was applied to the diplotypes in the patient population.

### Analysis of genotype count data in six cancers

The genome-wide genotype count data for the six cancer types were obtained from the website of the National Bioscience Database Center (NBDC) Human Database (http://humandbs.biosciencedbc.jp/). Each dataset had been quality controlled with sample call rate ≥ 0.98, SNP call rate ≥ 0.95 and p-value of original SNP-level HWT ≥ 1 x 10^-6^, and consisted of approximately 190 individuals. LD information was obtained using LDmatrix function of R package LDlinkR (34) from EAS population in the 1000 Genomes Phase 3 dataset (21). The variants overlapping with genes (within 2 kb upstream or downstream of the transcripts), which were determined using SNPnexus (52), and those with MAF ≥5% were selected. For esophageal, lung, breast, gastric, colorectal, and prostate cancers, we applied gene-HWT to 13,482 genes with 174,270 variants, 12,051 genes with 100,496 variants, 11,813 genes with 94,412 variants, 11,992 genes with 99,748 variants, 11,855 genes with 92,809 variants, and 11,857 genes with 92,690 variants, respectively. q-value (53), an FDR-adjusted p-value, was calculated using the q-value package in R. We combined *z*_*gene*_ values from the six cancers using Stouffer’s method. Specifically, the combined z-score, *z*_*gene*(*comb*)_, was computed by summing up the individual z-scores, *z*_*gene*(*i*)_, and dividing by the square root of the total number of studies, *k*( *=* 6):

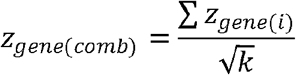

P-values (and subsequently q-values) were calculated based on the fact that, under the null hypothesis, *z*_*gene*(*comb*)_ follows the standard normal distribution.

### Statistics and bioinformatics tools

The following tools were used:

qvalue package in R: http://www.bioconductor.org/packages/release/bioc/html/qvalue.html

SNPnexus: https://www.snp-nexus.org/v4/

Hapsim package in R: https://cran.r-project.org/web/packages/hapsim/index.html

LDlinkR package in R: https://cran.r-project.org/web/packages/LDlinkR/index.html

## Data Availability

Genotypes from 1KG are available at http://ftp.ensembl.org/pub/data_files/homo_sapiens/GRCh38/variation_genotype/ALL.chr20_GRCh38.genotypes.20170504.vcf.gz.
Genotype counts data of six cancer types used for this research are available at the website of the NBDC Human Database / the Japan Science and Technology Agency (JST) (http://humandbs.biosciencedbc.jp/) through the following six accession numbers: hum0014.v2.jsnp.cc.v1, hum0014.v2.jsnp.pc.v1, hum0014.v2.jsnp.sc.v1, hum0014.v2.jsnp.bc.v1, hum0014.v2.jsnp.lc.v1, and hum0014.v2.jsnp.182ec.v1.
The R code for implementing gene-HWT is available at
https://github.com/jonishino/gene-HWT.git

http://humandbs.biosciencedbc.jp/

https://github.com/jonishino/gene-HWT.git

## DECLARATIONS

### Ethics approval and consent to participate

Not applicable.

### Consent for publication

Not applicable.

### Availability of data and materials

Genotypes from 1KG are available at http://ftp.ensembl.org/pub/data_files/homo_sapiens/GRCh38/variation_genotype/ALL.chr20_GRCh38.genotypes.20170504.vcf.gz.

Genotype counts data of six cancer types used for this research are available at the website of the NBDC Human Database / the Japan Science and Technology Agency (JST) (http://humandbs.biosciencedbc.jp/) through the following six accession numbers: hum0014.v2.jsnp.cc.v1, hum0014.v2.jsnp.pc.v1, hum0014.v2.jsnp.sc.v1, hum0014.v2.jsnp.bc.v1, hum0014.v2.jsnp.lc.v1, and hum0014.v2.jsnp.182ec.v1.

The R code for implementing gene-HWT is available at https://github.com/jonishino/gene-HWT.git

### Authors’ contributions

J.N. conceptualized and developed the methodology. J.N. performed the simulations and real data analysis, and wrote the manuscript. M.K., and F.M. contributed to the interpretation of results and discussions. All authors read and approved the final manuscript.

