## Supplementary Note for "Gene-based Hardy–Weinberg equilibrium test using genotype count data identifies novel cancer-related genes"

Derivation of $Cov(z_i, z_j) = r_{i,j}^2$

### 1. Setting

For the first locus on a gene with two alleles $a$ and $A$ with population frequencies $p$ and $q = 1 - p$, the statistic to test for Hardy-Weinberg equilibrium, z, in $n$ diploid samples is the following:

$$z = \frac{x_h - 2n\hat{p}\hat{q}}{2\hat{p}\hat{q}\sqrt{n}},$$

where $x_h$ is observed number of heterozygosity in the sample, and $\hat{p}$ and $\hat{q}$ represent the sample frequency of $a$ and $A$, respectively. For the second locus on the same genes, we use the same notations as the first locus, but with dash, i.e., two alleles $a'$ and $A'$, along with their population frequencies $p'$ and $q' = 1 - p'$, their corresponding sample frequencies $\hat{p}'$ and $\hat{q}'$, the statistics HWT $z'$ and observed number of heterozygosity $x_h{}'$. In the following, we prove that $Cov(z, z') = r^2$, and in the text, $Cov(z_i, z_j) = r_{i,j}^2$.

Neglecting the sampling variance for the denominator of $z$ and $z'$, the covariance between $z$ and $z'$ is expressed as,

$$Cov(z, z') = \frac{Cov(x_h - 2n\hat{p}\hat{q},\ x_h{}' - 2n\hat{p}'\hat{q}')}{4pqp'q'\mathrm{n}}$$

$$= \frac{Cov(x_h, x_h{}') - 2nCov(x_h, \hat{p}'\hat{q}') - 2nCov(\hat{p}\hat{q}, x_h') + 4n^2Cov(\hat{p}\hat{q}, \hat{p}'\hat{q}')}{4pqp'q'\mathrm{n}}.$$

In the following, we derive the covariances $Cov(x_h, x_h{}')$, $Cov(x_h, \hat{p}'\hat{q}')$, $Cov(\hat{p}\hat{q}, x_h')$ and $Cov(\hat{p}\hat{q}, \hat{p}'\hat{q}')$ of the numerator of $Cov(z, z')$ in terms of population parameters, i.e., allele frequencies and linkage disequilibrium coefficient between the two loci, $D$.

The population frequencies (probabilities) of haplotypes $aa'$, $aA'$, $Aa'$ and $AA'$ are given by $\mathrm{P}(aa') = pp' + D$, $\mathrm{P}(aA') = pq' - D$, $\mathrm{P}(Aa') = qp' - D$ and $\mathrm{P}(AA') = qq' + D$, respectively. A diplotype of one sample is determined by sampling the haplotypes according to their frequencies with replacement from the population, independently of the other diplotypes. Index variable $h_j$ indicates whether the $j$-th sample is heterozygous for the first locus. The $j$-th sample is composed of the $(2j$-$1)$-th and $2j$-th chromosomes: the 1st and 2nd chromosomes belong to the 1st sample, the 3rd and 4th chromosomes belong to the 2nd sample, and so on. Furthermore, let $a_1, a_2, a_3, \ldots, a_{2n}$ be the index variables indicating whether the 1st, 2nd, ..., $2n$-th chromosomes carry the allele "$a$", and let $A_1, A_2, A_3, \ldots, A_{2n}$

indicates whether the 1st, 2nd, ..., $2n$-th chromosomes carry the allele "$A$". For the second locus, the same symbols with dashes are used, e.g., $h_2{}'$, $a'_4$, $A'_1$.

### 2. $Cov(x_h, x_h{}')$

$Cov(x_h, x_h{}')$ is simplified as

$$\begin{aligned}&Cov(x_h, x_h{}')\\&= Cov(h_1 + \cdots + h_n, h'_1 + \cdots + h'_n)\\&= Cov(h_1, h'_1) + Cov(h_2, h'_2) + \cdots + Cov(h_n, h'_n)\\&= nCov(h_1, h'_1),\end{aligned}$$

since $Cov(h_i, h'_j) = 0$ $(i \neq j)$.

Considering $h_1 = a_1A_2 + A_1a_2$ and $h'_1 = a'_1A'_2 + A'_1a'_2$,

$$\begin{aligned}&Cov(x_h, x_h{}')\\&= nCov(a_1A_2 + A_1a_2, a'_1A'_2 + A'_1a'_2)\\&= n\{Cov(a_1A_2, a'_1A'_2) + Cov(a_1A_2, A'_1a'_2) + Cov(A_1a_2, a'_1A'_2) + Cov(A_1a_2, A'_1a'_2)\}\end{aligned}$$

Here,

$$\begin{aligned}Cov(a_1A_2, a'_1A'_2) &= E(a_1A_2a'_1A'_2) - E(a_1A_2)E(a'_1A'_2)\\&= E(a_1a'_1)E(A_2A'_2) - E(a_1)E(A_2)E(a'_1)E(A'_2)\\&= (pp' + D)(qq' + D) - pqp'q' = (pp' + qq')D + D^2.\end{aligned}$$

Similarly we get

$$\begin{aligned}Cov(a_1A_2, A'_1a'_2) &= E(a_1A'_1)E(A_2a'_2) - E(a_1)E(A_2)E(A'_1)E(a'_2)\\&= (pq' - D)(qp' - D) - pqp'q' = -(pq' + qp')D + D^2\\Cov(A_1a_2, a'_1A'_2) &= E(A_1a'_1)E(a_2A'_2) - E(A_1)E(a_2)E(a'_1)E(A'_2)\\&= (qp' - D)(pq' - D) - pqp'q' = -(pq' + qp')D + D^2\\Cov(A_1a_2, A'_1a'_2) &= E(A_1A'_1)E(a_2a'_2) - E(a_1)E(A_2)E(a'_1)E(A'_2)\\&= (qq' + D)(pp' + D) - pqp'q' = (pp' + qq')D + D^2.\end{aligned}$$

Then,

$$\begin{aligned}&Cov(x_h, x_h{}')\\&= n\{(pp' + qq')D + D^2 - (pq' + qp')D + D^2 - (pq' + qp')D + D^2 + (pp' + qq')D + D^2\}\\&= 2n(2p - 1)(2p' - 1)D + 4nD^2.\end{aligned}$$

### 3. $Cov(x_h, \hat{p}'\hat{q}')$ and $Cov(\hat{p}\hat{q}, x'_h)$

$Cov(x_h, \hat{p}'\hat{q}')$ is be expressed using $4n^3$ simpler covariances, $Cov(h_., a'_.A'_.)$'s, as,

$$Cov(x_h, \hat{p}'\hat{q}') = Cov\left( (h_1 + \cdots + h_n), \frac{(a'_1 + \cdots + a'_{2n})}{2n} \frac{(A'_1 + \cdots + A'_{2n})}{2n} \right)$$

$$= \frac{1}{4n^2} Cov\big((h_1 + \cdots + h_n), (a'_1 + \cdots + a'_{2n})(A'_1 + \cdots + A'_{2n})\big)$$

$$= \frac{1}{4n^2} \{Cov(h_1, a'_1 A'_1) + Cov(h_1, a'_1 A'_2) + \cdots + Cov(h_n, a'_{2n} A'_{2n})\}.$$

Many covariances are zero. For example, $cov(h_1, a'_1 A'_1) = 0$ since $a'_1 A'_1 = 0$. $Cov(h_1, a'_3 A'_4)$ is also 0 since $h_1$ and $a'_3 A'_4$ are independent. $Cov(x_h, \hat{p}'\hat{q}')$ can be expressed using three types of non-zero covariances with the same values. One type is that $a'_.$ and $A'_.$ refer different chromosomes but in the same sample that $h_.$ refers, another is $a'_.$ refers one chromosome in a sample that $h_.$ refers but $A'$ do not, and the last one is $A'_.$ refers one chromosome in a sample that $h_.$ refers but $a'$ do not, which appear $2n$ times, $4n(n-1)$ times, and $4n(n-1)$ times, respectively. Then,

$$Cov(x_h, \hat{p}'\hat{q}') =$$

$$= \frac{1}{4n^2} \{2nCov(h_1, a'_1 A'_2) + 4n(n-1)\big(Cov(h_1, a'_1 A'_3) + Cov(h_1, A'_1 a'_3)\big)\}$$

$$= \frac{1}{2n} Cov(h_1, a'_1 A'_2) + \frac{(n-1)}{n} \big(Cov(h_1, a'_1 A'_3) + Cov(h_1, A'_1 a'_3)\big).$$

Considering $h_1 = a_1 A_2 + A_1 a_2$,

$$Cov(x_h, \hat{p}'\hat{q}')$$

$$= \frac{1}{2n} \{Cov(a_1 A_2, a'_1 A'_2) + Cov(A_1 a_2, a'_1 A'_2)\}$$

$$+ \frac{(n-1)}{n} \{Cov(a_1 A_2, a'_1 A'_3) + Cov(A_1 a_2, a'_1 A'_3) + Cov(a_1 A_2, A'_1 a'_3)$$

$$+ Cov(A_1 a_2, A'_1 a'_3)\}.$$

Here, $Cov(a_1 A_2, a'_1 A'_2)$ and $Cov(A_1 a_2, a'_1 A'_2)$ are as mentioned earlier. Other covariances are as follows.

$$Cov(a_1 A_2, a'_1 A'_3) = E(a_1 A_2 a'_1 A'_3) - E(a_1 A_2) E(a'_1 A'_3)$$

$$= E(a_1 a'_1) E(A_2) E(A'_3) - E(a_1) E(A_2) E(a'_1) E(A'_3) = (pp' + D) qq' - pqp'q'$$

$$= qq'D,$$

$$Cov(A_1 a_2, a'_1 A'_3) = E(A_1 a_2 a'_1 A'_3) - E(A_1 a_2) E(a'_1 A'_3)$$

$$= E(A_1 a'_1) E(a_2) E(A'_3) - E(A_1) E(a_2) E(a'_1) E(A'_3) = (qp' - D) pq' - pqp'q'$$

$$= -pq'D,$$

$$Cov(a_1A_2, A'_1a'_3) = E(a_1A_2, A'_1a'_3) - E(a_1A_2)E(A'_1a'_3)$$
$$= E(a_1A'_1)E(A_2)E(a'_3) - E(A_1)E(a_2)E(a'_1)E(A'_3) = (pq' - D)qp' - pqp'q'$$
$$= -qp'D \text{ and}$$
$$Cov(A_1a_2, A'_1a'_3) = E(A_1a_2A'_1a'_3) - E(A_1a_2)E(A'_1a'_3)$$
$$= E(A_1A'_1)E(a_2)E(a'_3) - E(A_1)E(a_2)E(A'_1)E(a'_3) = (qq' + D)pp' - pqp'q'$$
$$= pp'D.$$

Then we get

$$Cov(x_h, \hat{p}'\hat{q}')$$
$$= \frac{1}{2n}\{(pp' + qq')D + D^2 - (pq' + qp')D + D^2\} + \frac{(n-1)}{n}\{qq'D - pq'D - qp'D + pp'D\}$$
$$= \frac{1}{2n}\{(2p-1)(2p'-1)D + 2D^2\} + \frac{(n-1)}{n}\{(2p-1)(2p'-1)D\}$$
$$= \frac{(2n-1)(2p-1)(2p'-1)D + 2D^2}{2n}.$$

This is symmetrical with respect to the two loci, then $Cov(\hat{p}\hat{q}, x'_h) = Cov(x_h, \hat{p}'\hat{q}')$.

4. $Cov(\hat{p}\hat{q}, \hat{p}'\hat{q}')$

$Cov(\hat{p}\hat{q}, \hat{p}'\hat{q}')$ is expressed using $(4n)^4$ covariances, $Cov(a.A., a'.A'.)$'s, as

$$Cov(\hat{p}\hat{q}, \hat{p}'\hat{q}') = Cov\left(\frac{(a_1 + \cdots + a_{2n})}{2n}\frac{(A_1 + \cdots + A_{2n})}{2n}, \frac{(a'_1 + \cdots + a'_{2n})}{2n}\frac{(A'_1 + \cdots + A'_{2n})}{2n}\right)$$
$$= \frac{1}{16n^4}Cov\big((a_1 + \cdots + a_{2n})(A_1 + \cdots + A_{2n}), (a'_1 + \cdots + a'_{2n})(A'_1 + \cdots + A'_{2n})\big)$$
$$= \frac{1}{16n^4}\{Cov(a_1A_1, a'_1A'_1) + Cov(a_1A_1, a'_1A'_2) + \cdots + Cov(a_{2n}A_{2n}, a'_{2n}A'_{2n})\}.$$

The covariances, $Cov(a.A., a'.A'.)$'s, can be categorized into 16 types. The classification of $Cov(a.A., a'.A'.)$'s and their values (the derivations already mentioned) and appearances are shown in the table below. There are six types of non-zero covariances.

From the table,

$Cov(\hat{p}\hat{q}, \hat{p}'\hat{q}') =$

$$= \frac{1}{16n^4}[2n(2n-1)\{Cov(a_1A_2, a'_1A'_2) + Cov(a_1A_2, A'_1a'_2)\}$$
$$+ 2n(2n-1)(2n-2)\{Cov(a_1A_2, a'_1A'_3) + Cov(A_1a_2, a'_1A'_3)$$
$$+ Cov(a_1A_2, A'_1a'_3) + Cov(A_1a_2, A'_1a'_3)\}]$$

$$= \frac{1}{16n^4}[2n(2n-1)\{(2p-1)(2p'-1)D + 2D^2\} + 2n(2n-1)(2n-2)\{(2p-1)(2p'-1)D\}]$$

$$= \frac{1}{8n^3}[(2n-1)^2(2p-1)(2p'-1)D + 2(2n-1)D^2].$$

Classification of $Cov(a.A., a'.A'.)$'s that constitute $Cov(\hat{p}\hat{q}, \hat{p}'\hat{q}')$.

| Chromosomes referred by $a., A., a'., A'.$ | Type | Example | Appearance | Value of covariance |
|---|---|---|---|---|
| One chromosome | $a.$, $A.$, $a'.$, $A'.$ refers one chromosome | $Cov(a_1A_1, a'_1A'_1)$ | $2n$ | 0 |
| One chromosome by 3 variables, another by 1. | All except $A'.$ refer one chromosome. | $Cov(a_1A_1, a'_1A'_2)$ | $2n(2n-1)$ | 0 |
| | All except $a'.$ refer one chromosome. | $Cov(a_1A_1, A'_1a'_2)$ | 〃 | 0 |
| | All except $A.$ refer one chromosome. | $Cov(a_1A_2, a'_1A'_1)$ | 〃 | 0 |
| | All except $a.$ refer one chromosome. | $Cov(A_1a_2, a'_1A'_1)$ | 〃 | 0 |
| One chromosome by 2 variables, another by 2. | $a.$ and $A.$ ($a'.$ and $A'.$) refer one chromosome. | $Cov(a_1A_1, a'_2A'_2)$ | $2n(2n-1)$ | 0 |
| | $a.$ and $A'.$ ($A.$ and $a'.$) refer one chromosome. | $Cov(a_1A_2, A'_1a'_2)$ | 〃 | $-(pq' + qp')D + D^2$ |
| | $a.$ and $a'.$ ($A.$ and $A'.$) refer one chromosome. | $Cov(a_1A_2, a'_1A'_2)$ | 〃 | $(pp' + qq')D + D^2$ |
| One chromosome by 2 variables, another by 1, and | $a.$ and $A.$ refer one chromosome. $a'.$ and $A'.$ each refer | $Cov(a_1A_1, a'_2A'_3)$ | $2n(2n-1)(2n-2)$ | 0 |

| | | | | |
|---|---|---|---|---|
| yet another by 1. | different chromosomes. | | | |
| | $a'$. and $A'$. refer one chromosome. $a$. and $A$. each refer different chromosomes. | $Cov(a_2A_3, a'_1A'_1)$ | 〃 | 0 |
| | $a$. and $a'$. refer one chromosome. $A$. and $A'$. each refer different chromosomes. | $Cov(a_1A_2, a'_1A'_3)$ | 〃 | $qq'D$ |
| | $a$. and $A'$. refer one chromosome. $A$. and $a'$. each refer different chromosomes. | $Cov(a_1A_2, A'_1a'_3)$ | 〃 | $-qp'D$ |
| | $A$. and $a'$. refer one chromosome. $a$. and $A'$. each refer different chromosomes. | $Cov(A_1a_2, a'_1A'_3)$ | 〃 | $-pq'D$ |
| | $A$. and $A'$. refer one chromosome. $a$. and $a'$. each refer different chromosomes. | $Cov(A_1a_2, A'_1a'_3)$ | 〃 | $pp'D$ |
| Four chromosomes by 4 variables. | $a$., $A$., $a'$., and $A'$. each refer different chromosomes. | $Cov(A_1a_2, A'_3a'_4)$ | $2n(2n-1)(2n-2)(2n-3)$ | 0 |
| - | - | - | Total: $16n^4$ | - |

### 5. $Cov(z, z')$

Revisiting $Cov(z, z')$,

$$Cov(z, z') = \frac{Cov(x_h, x_h') - 2nCov(x_h, \hat{p}'\hat{q}') - 2nCov(\hat{p}\hat{q}, x_h') + 4n^2Cov(\hat{p}\hat{q}, \hat{p}'\hat{q}')}{4pqp'q'n}$$

$$= \frac{Cov(x_h, x_h') - 4nCov(x_h, \hat{p}'\hat{q}') + 4n^2Cov(\hat{p}\hat{q}, \hat{p}'\hat{q}')}{4pqp'q'n}$$

From the above,

$$Cov(x_h, x_h{}') = 2n(2p-1)(2p'-1)D + 4nD^2,$$

$$Cov(x_h, \hat{p}'\hat{q}') = \frac{(2n-1)(2p-1)(2p'-1)D + 2D^2}{2n} \text{ and}$$

$$Cov(\hat{p}\hat{q}, \hat{p}'\hat{q}') = \frac{1}{8n^3}[(2n-1)^2(2p-1)(2p'-1)D + 2(2n-1)D^2].$$

Substituting these into the expression for $Cov(z, z')$, we can simplify to,

$$Cov(z, z') = \frac{(2p-1)(2p'-1)}{8n^2}\frac{r}{\sqrt{pqp'q'}} + \frac{4n^2-2n-1}{4n^2}r^2$$

As $n$ becomes large $Cov(z, z')$ approaches to $r^2$.
