## Supplementary Table 1 for "Gene-based Hardy–Weinberg equilibrium test using genotype count data identifies novel cancer-related genes"

Supplementary Table 1. Genes identified by gene-HWT with *q*-value <0.2 for each cancer type.

| Cancer | Gene | # of SNPs | gene-HWT | | |
|---|---|---|---|---|---|
| | | | $Z_{gene}$ | *p*-value | *q*-value |
| esophageal | BRAP | 2 | 5.98 | 2.20E-09 | 2.96E-05 |
| esophageal | CCDC9 | 2 | -4.19 | 2.75E-05 | 1.60E-01 |
| esophageal | CRB3 | 2 | -4.08 | 4.49E-05 | 1.60E-01 |
| esophageal | GPR39 | 50 | -4.07 | 4.74E-05 | 1.60E-01 |
| lung | HLA-DMA | 1 | -4.65 | 3.31E-06 | 2.40E-02 |
| lung | APOBEC3A | 1 | -4.59 | 4.51E-06 | 2.40E-02 |
| lung | OR4N2 | 5 | 4.53 | 5.97E-06 | 2.40E-02 |
| lung | GYG1 | 2 | -4.44 | 9.10E-06 | 2.74E-02 |
| lung | ZCWPW1 | 1 | 4.32 | 1.55E-05 | 3.73E-02 |
| lung | QRFPR | 9 | -4.24 | 2.21E-05 | 4.36E-02 |
| lung | SELENOV | 1 | 4.20 | 2.69E-05 | 4.36E-02 |
| lung | AL163636.2 | 1 | -4.18 | 2.89E-05 | 4.36E-02 |
| lung | MAFK | 1 | 4.15 | 3.27E-05 | 4.37E-02 |
| lung | VIM | 1 | 4.06 | 4.93E-05 | 5.94E-02 |
| lung | CD40 | 1 | 3.93 | 8.57E-05 | 9.38E-02 |
| lung | SETMAR | 3 | -3.83 | 1.28E-04 | 1.28E-01 |
| lung | SH3GLB2 | 1 | 3.79 | 1.50E-04 | 1.39E-01 |
| lung | C15orf53 | 35 | -3.73 | 1.91E-04 | 1.59E-01 |
| lung | ZNF736 | 1 | -3.71 | 2.09E-04 | 1.59E-01 |
| lung | AOX1 | 21 | -3.71 | 2.11E-04 | 1.59E-01 |
| lung | CCSER2 | 11 | -3.68 | 2.34E-04 | 1.66E-01 |
| lung | ZNF680 | 2 | -3.65 | 2.58E-04 | 1.72E-01 |
| lung | PDLIM3 | 2 | -3.61 | 3.01E-04 | 1.91E-01 |
| breast | OR4N2 | 2 | 4.82 | 1.45E-06 | 1.71E-02 |
| breast | ZNF736 | 1 | -4.65 | 3.26E-06 | 1.92E-02 |
| breast | CTSO | 2 | -4.44 | 9.09E-06 | 3.58E-02 |
| breast | ZNF680 | 1 | -4.28 | 1.85E-05 | 4.69E-02 |
| breast | KIAA2013 | 1 | -4.27 | 1.99E-05 | 4.69E-02 |
| breast | HLA-DMA | 1 | -4.18 | 2.91E-05 | 5.50E-02 |
| breast | HAVCR2 | 3 | -4.15 | 3.26E-05 | 5.50E-02 |
| breast | IGSF9B | 4 | -4.09 | 4.36E-05 | 6.44E-02 |
| breast | PLEKHG1 | 36 | -3.93 | 8.65E-05 | 1.14E-01 |
| breast | HRK | 4 | 3.81 | 1.39E-04 | 1.61E-01 |
| breast | PSMC1 | 1 | -3.79 | 1.50E-04 | 1.61E-01 |
| gastric | NUP54 | 1 | -5.14 | 2.75E-07 | 3.30E-03 |
| gastric | PRAMEF1 | 1 | -4.98 | 6.28E-07 | 3.77E-03 |
| gastric | RAD51 | 1 | -4.61 | 4.12E-06 | 1.65E-02 |
| gastric | PPP1CB | 3 | -4.43 | 9.24E-06 | 2.77E-02 |
| gastric | FKBP9 | 1 | 4.23 | 2.38E-05 | 5.42E-02 |
| gastric | CCR7 | 1 | -4.20 | 2.71E-05 | 5.42E-02 |
| gastric | KCNJ4 | 3 | -4.14 | 3.45E-05 | 5.92E-02 |
| gastric | RNGTT | 21 | -4.03 | 5.52E-05 | 8.28E-02 |
| gastric | ZFP14 | 1 | -3.94 | 8.24E-05 | 9.92E-02 |
| gastric | NMUR1 | 1 | -3.94 | 8.28E-05 | 9.92E-02 |
| gastric | CDR2 | 1 | -3.75 | 1.74E-04 | 1.75E-01 |
| gastric | KRT2 | 1 | -3.73 | 1.90E-04 | 1.75E-01 |
| gastric | GOLGA8A | 1 | -3.73 | 1.90E-04 | 1.75E-01 |
| gastric | TNFRSF10B | 3 | 3.69 | 2.24E-04 | 1.89E-01 |
| gastric | TBK1 | 1 | -3.68 | 2.37E-04 | 1.89E-01 |
| colorectal | OR14J1 | 1 | -5.11 | 3.16E-07 | 3.75E-03 |
| colorectal | DGKE | 2 | -4.87 | 1.12E-06 | 6.66E-03 |
| colorectal | ZNF701 | 1 | -4.74 | 2.13E-06 | 8.42E-03 |
| colorectal | ANO3 | 31 | -4.62 | 3.75E-06 | 9.89E-03 |
| colorectal | POFUT2 | 1 | -4.60 | 4.17E-06 | 9.89E-03 |
| colorectal | AL163636.2 | 1 | -4.33 | 1.47E-05 | 2.90E-02 |
| colorectal | TLX1 | 1 | 4.19 | 2.80E-05 | 4.74E-02 |

| Cancer | Gene | # of SNPs | gene-HWT | | |
|---|---|---|---|---|---|
| | | | $Z_{gene}$ | *p*-value | *q*-value |
| colorectal | DYNC1LI1 | 1 | −4.05 | 5.23E−05 | 7.75E−02 |
| colorectal | GOLIM4 | 5 | −3.91 | 9.32E−05 | 9.65E−02 |
| colorectal | TRDMT1 | 5 | −3.90 | 9.54E−05 | 9.65E−02 |
| colorectal | EML6 | 23 | −3.90 | 9.69E−05 | 9.65E−02 |
| colorectal | RAPGEF2 | 9 | −3.90 | 9.77E−05 | 9.65E−02 |
| colorectal | HSF2 | 5 | −3.86 | 1.13E−04 | 1.02E−01 |
| colorectal | FAM3C | 1 | −3.85 | 1.20E−04 | 1.02E−01 |
| colorectal | TP53 | 2 | 3.80 | 1.45E−04 | 1.11E−01 |
| colorectal | HLA-DMA | 1 | −3.79 | 1.50E−04 | 1.11E−01 |
| colorectal | RYR2 | 103 | −3.78 | 1.59E−04 | 1.11E−01 |
| colorectal | POPDC2 | 4 | −3.73 | 1.91E−04 | 1.26E−01 |
| colorectal | FSTL4 | 48 | −3.71 | 2.08E−04 | 1.30E−01 |
| colorectal | MRPL19 | 3 | −3.61 | 3.09E−04 | 1.83E−01 |
| prostate | NUP54 | 1 | −4.62 | 3.79E−06 | 4.50E−02 |
| prostate | ZNF680 | 1 | −4.40 | 1.07E−05 | 6.36E−02 |
| prostate | AL163636.2 | 1 | −4.21 | 2.56E−05 | 9.31E−02 |
| prostate | AC007998.2 | 2 | −4.16 | 3.14E−05 | 9.31E−02 |
| prostate | HLA-DMA | 1 | −4.10 | 4.18E−05 | 9.91E−02 |
| prostate | TBK1 | 1 | −4.03 | 5.70E−05 | 1.13E−01 |
| prostate | FBXO5 | 1 | −3.92 | 9.01E−05 | 1.37E−01 |
| prostate | FBXL4 | 7 | −3.91 | 9.27E−05 | 1.37E−01 |
| prostate | KLK11 | 1 | −3.84 | 1.21E−04 | 1.59E−01 |
| prostate | QRFPR | 11 | −3.79 | 1.50E−04 | 1.78E−01 |
| prostate | SRRM1 | 1 | 3.75 | 1.77E−04 | 1.91E−01 |
