## Supplementary Figure 1 for "Gene-based Hardy–Weinberg equilibrium test using genotype count data identifies novel cancer-related genes"

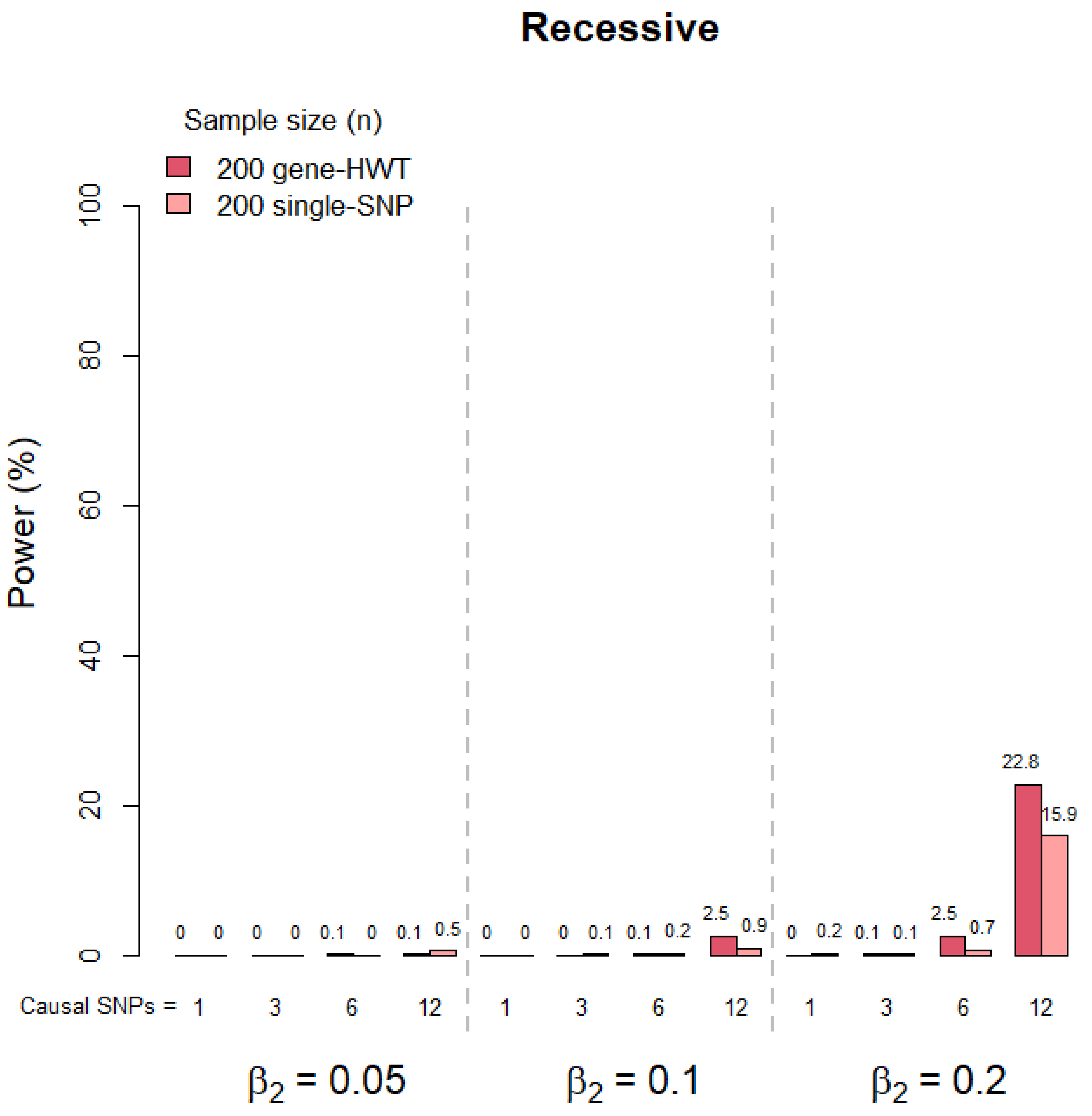


**Supplementary Figure 1. Comparison of power of gene-HWT with single SNP test.** Comparison of gene-HWT power with single SNP test at 0.05 significance, each with corrections. For 1000 genes (1000 simulations per parameter set), in gene-HWT Bonferroni correction was applied for the 1000 tests. In single-SNP test Bonferroni correction was applied for the number of SNPs
within each of 1000 genes (on average 78,537 SNPs).

Supplementary Figure 1. Countineud.

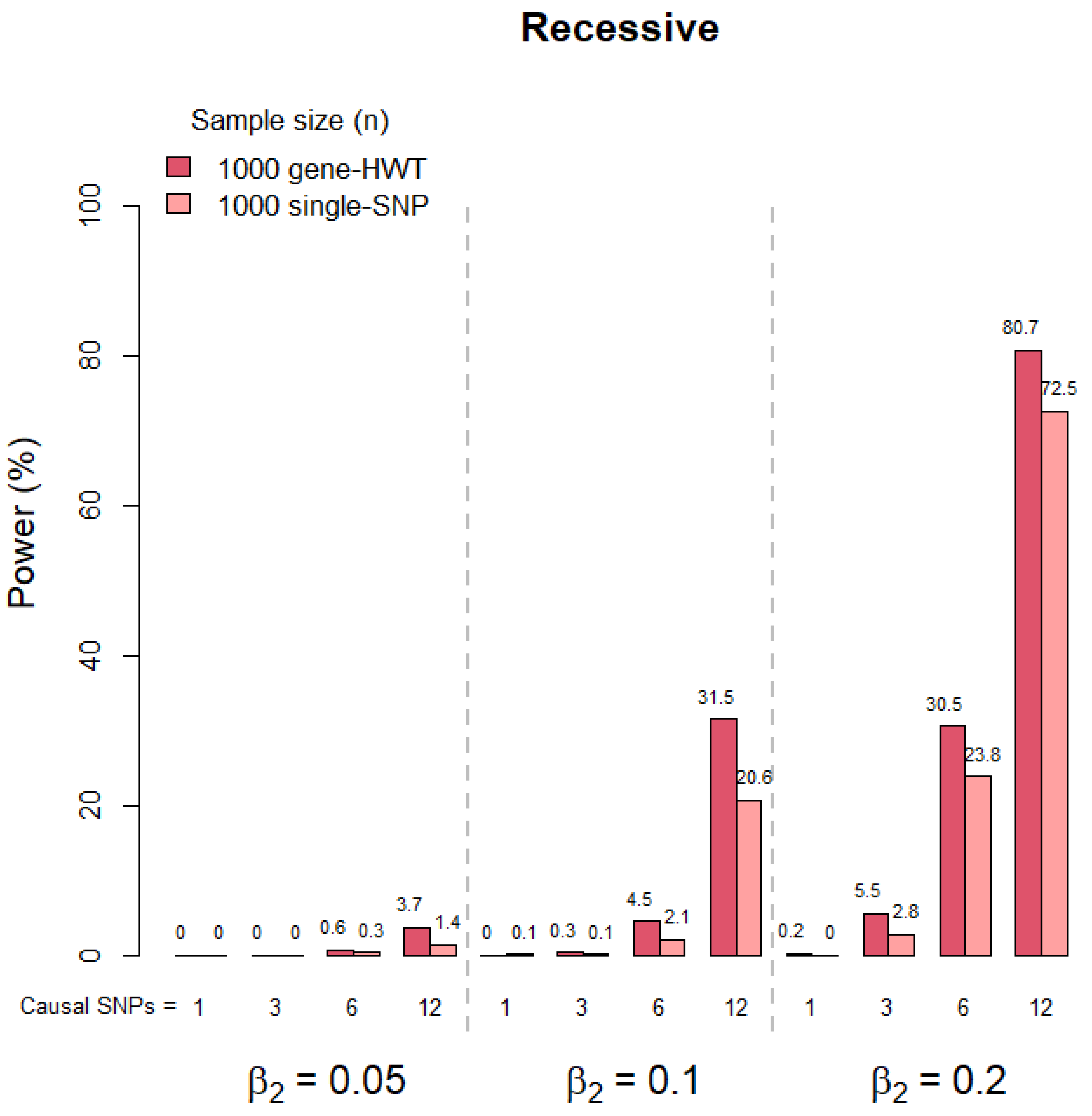

Supplementary Figure 1. Countineud.

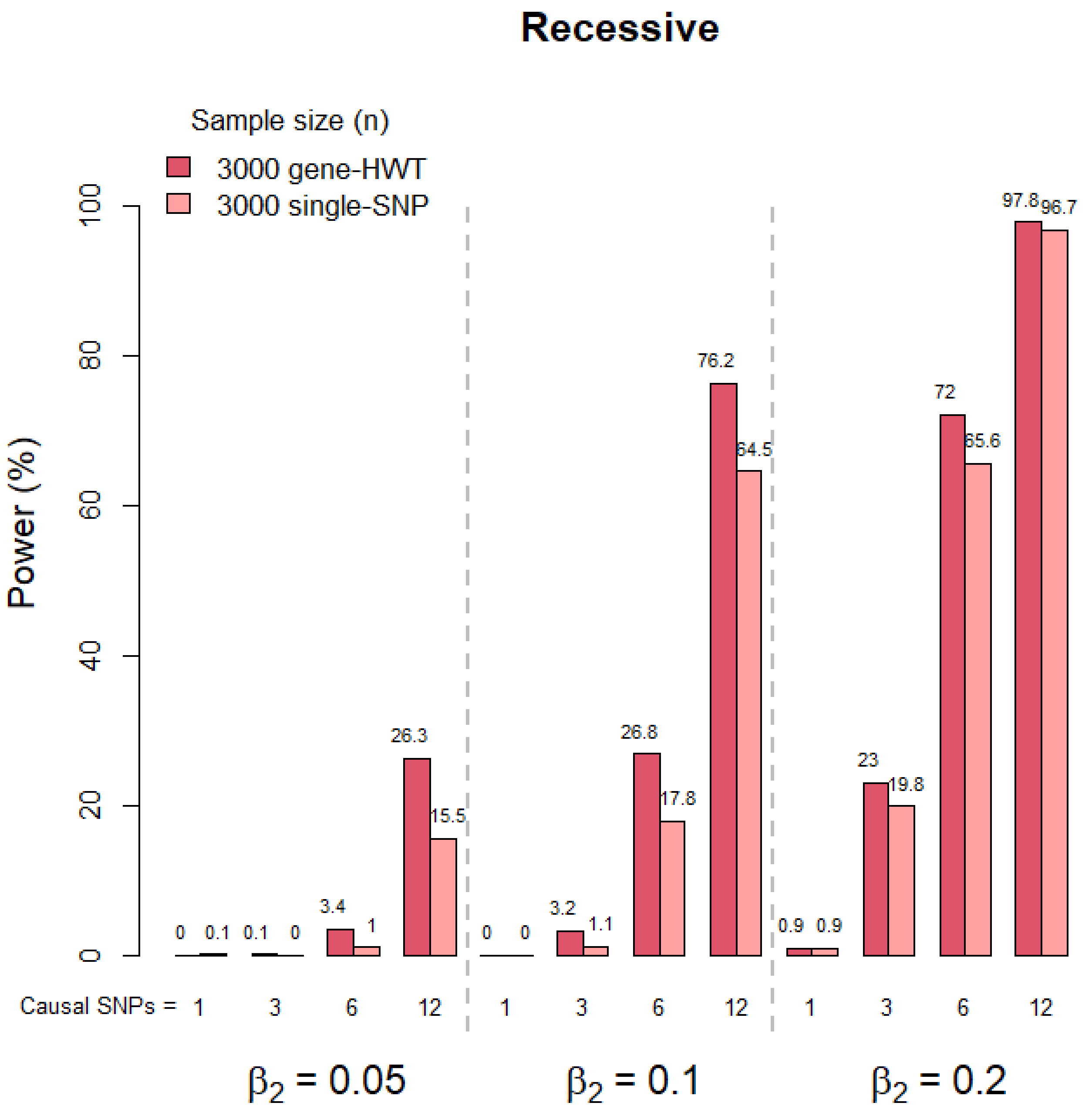

Supplementary Figure 1. Countineud.

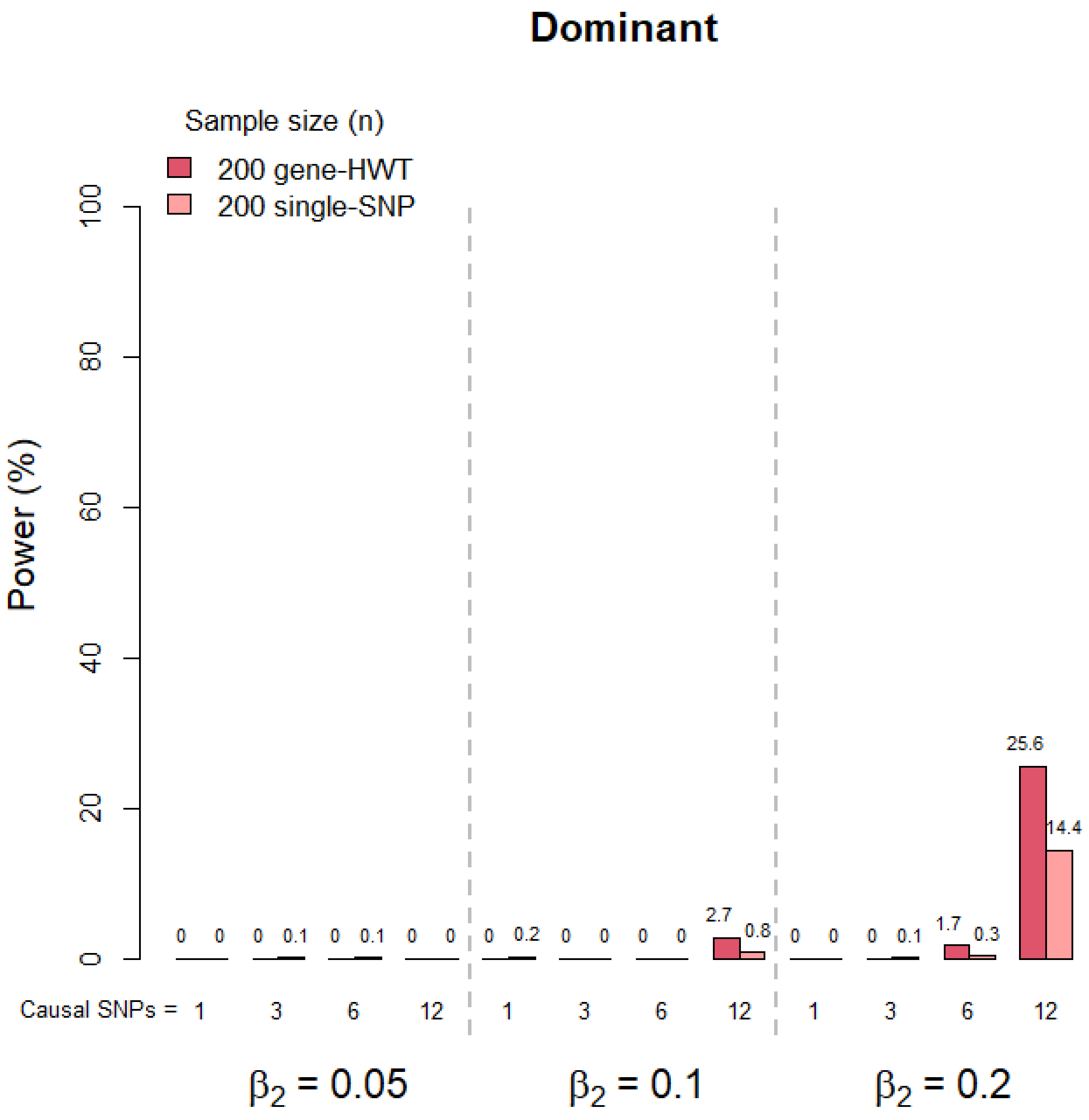

Supplementary Figure 1. Countineud.

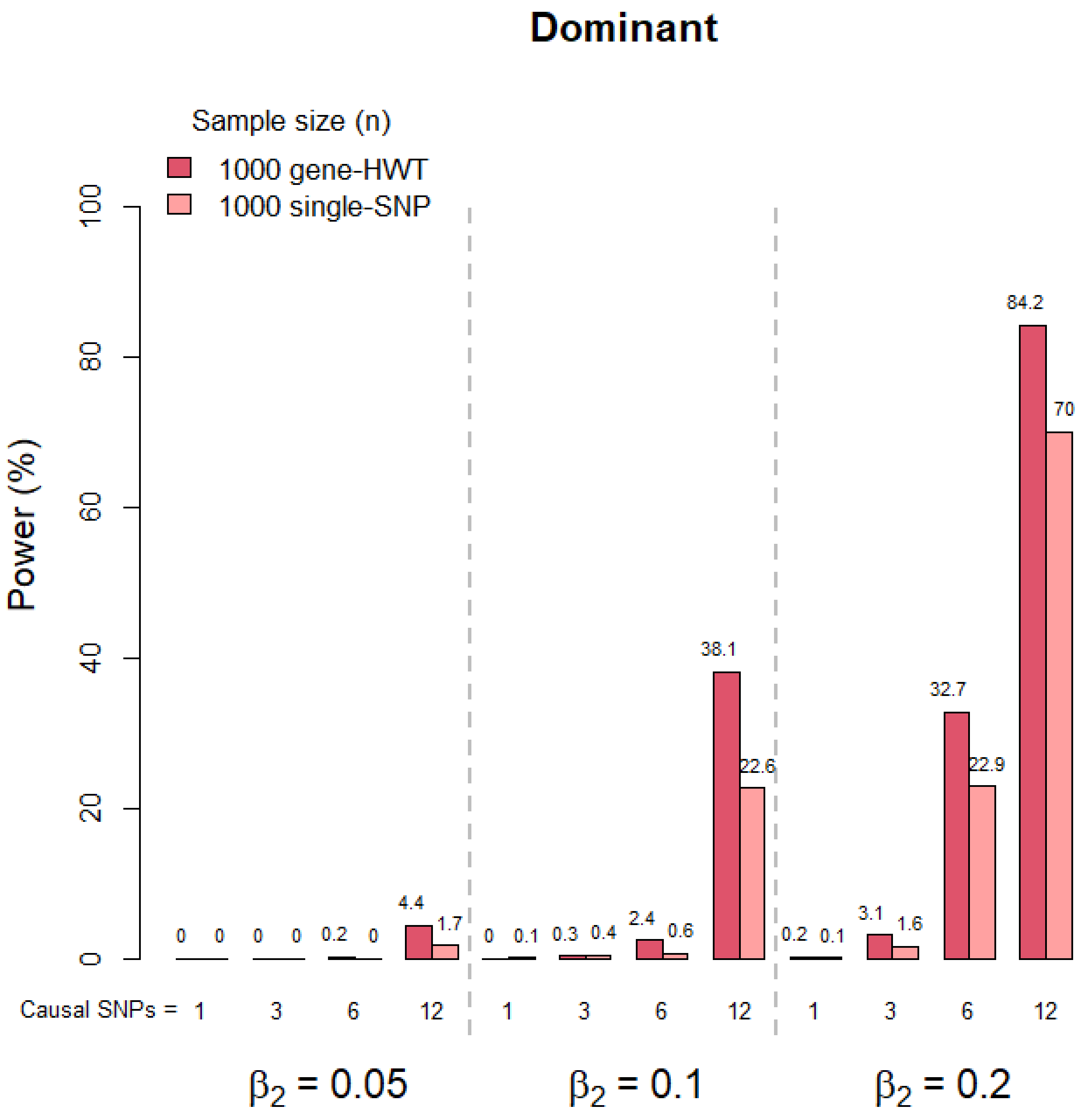

Supplementary Figure 1. Countineud.

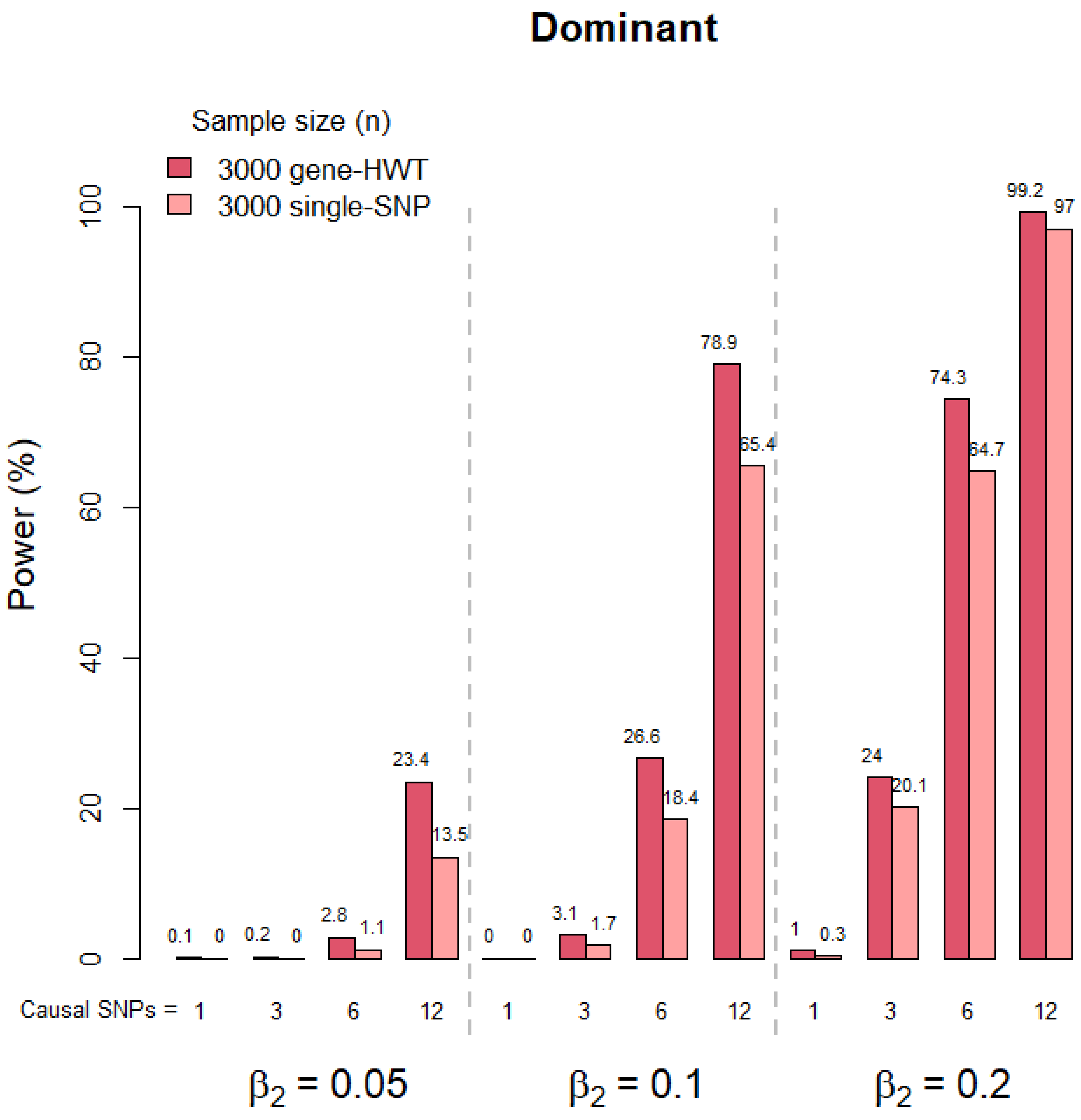

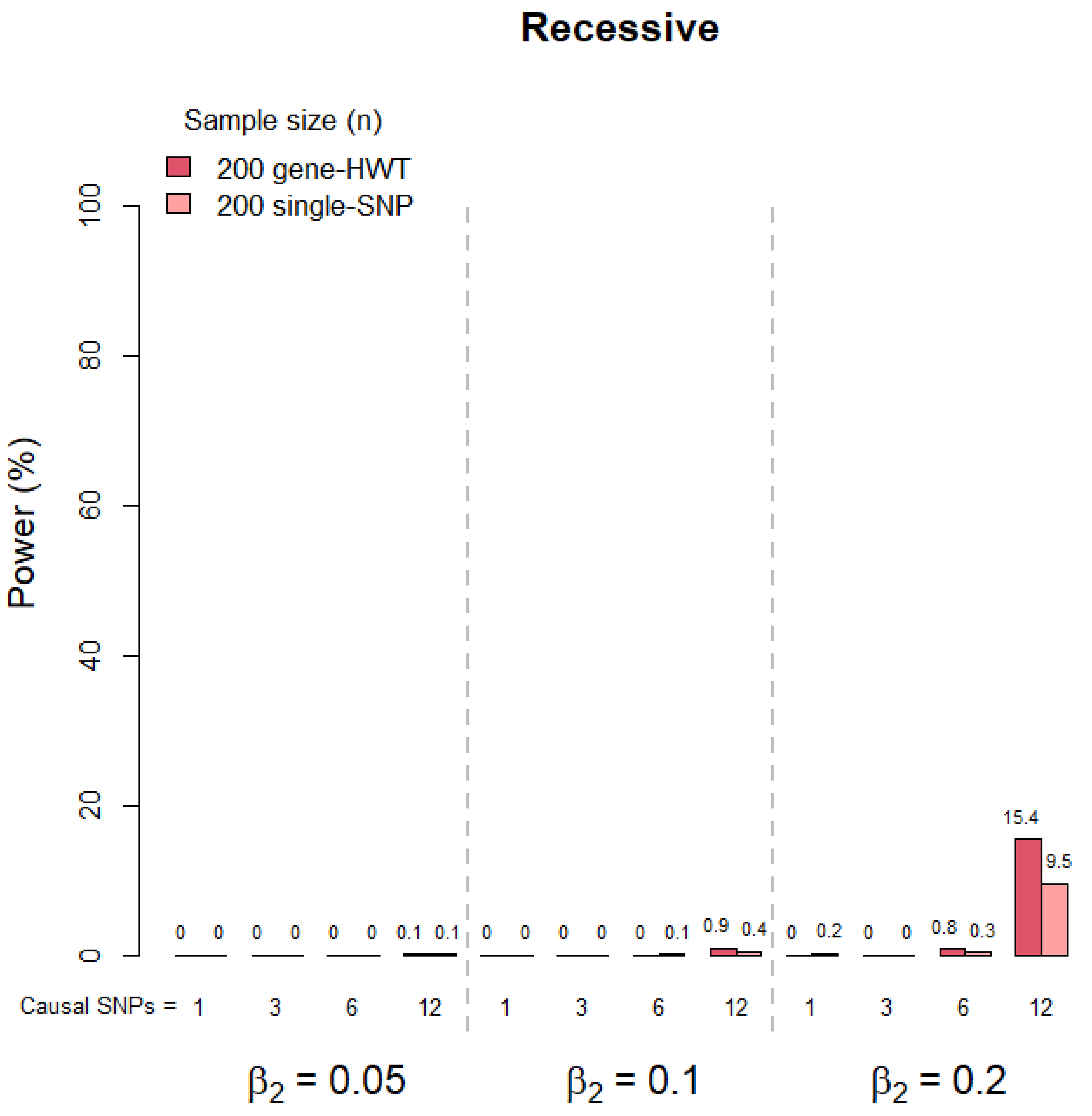


**Supplementary Figure 2. Comparison of power of gene-HWT with single- SNP test with the standard genome-wide significance levels.** Comparison of power of gene-HWT with single SNP test using commonly used genome-wide significance levels. GHWT used 20,000 genes (P < 2.5×10-6), single SNP test used 1 million SNPs (P < 5×10-8).

Supplementary Figure 2. Countineud.

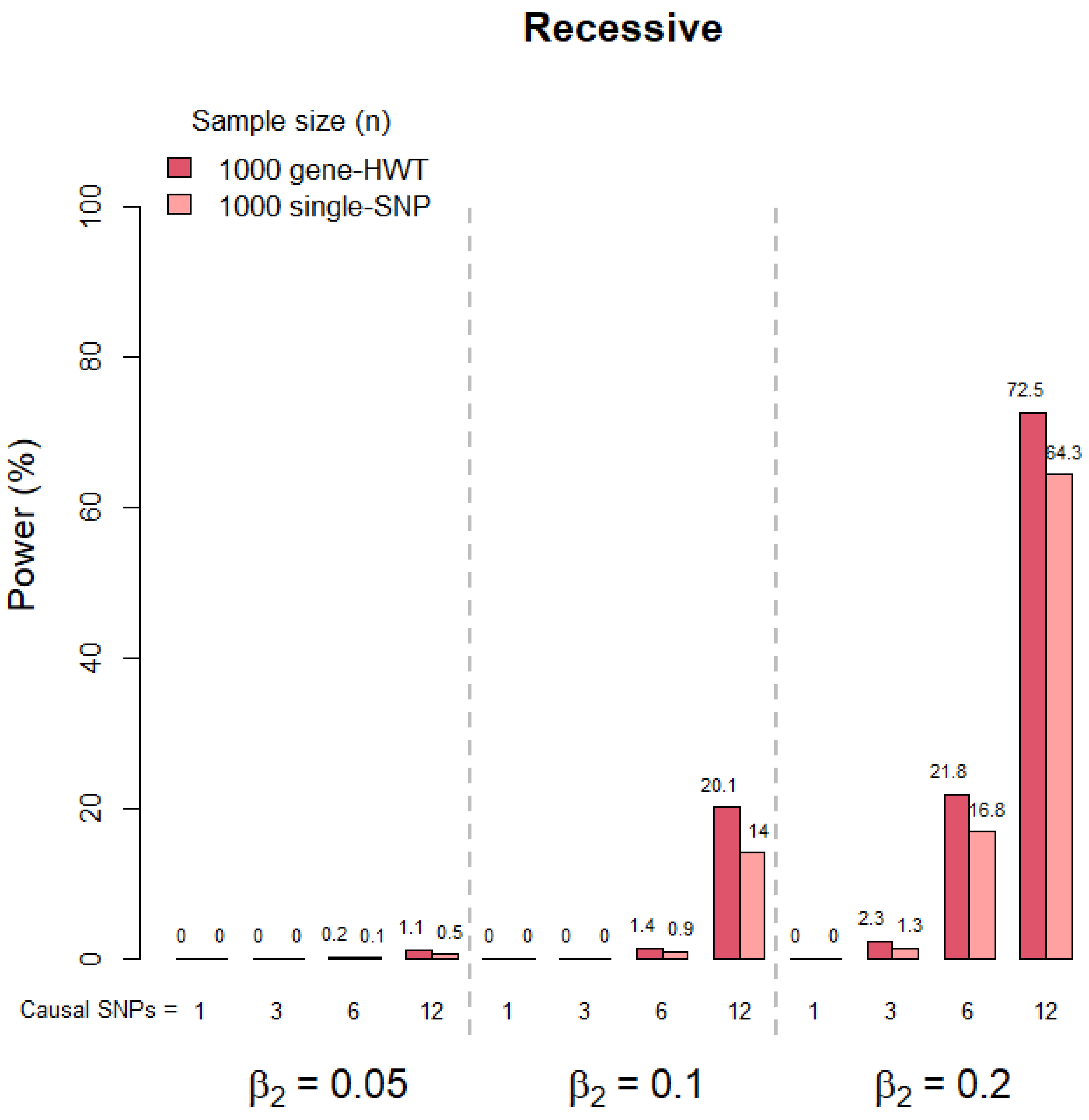

Supplementary Figure 2. Countineud.

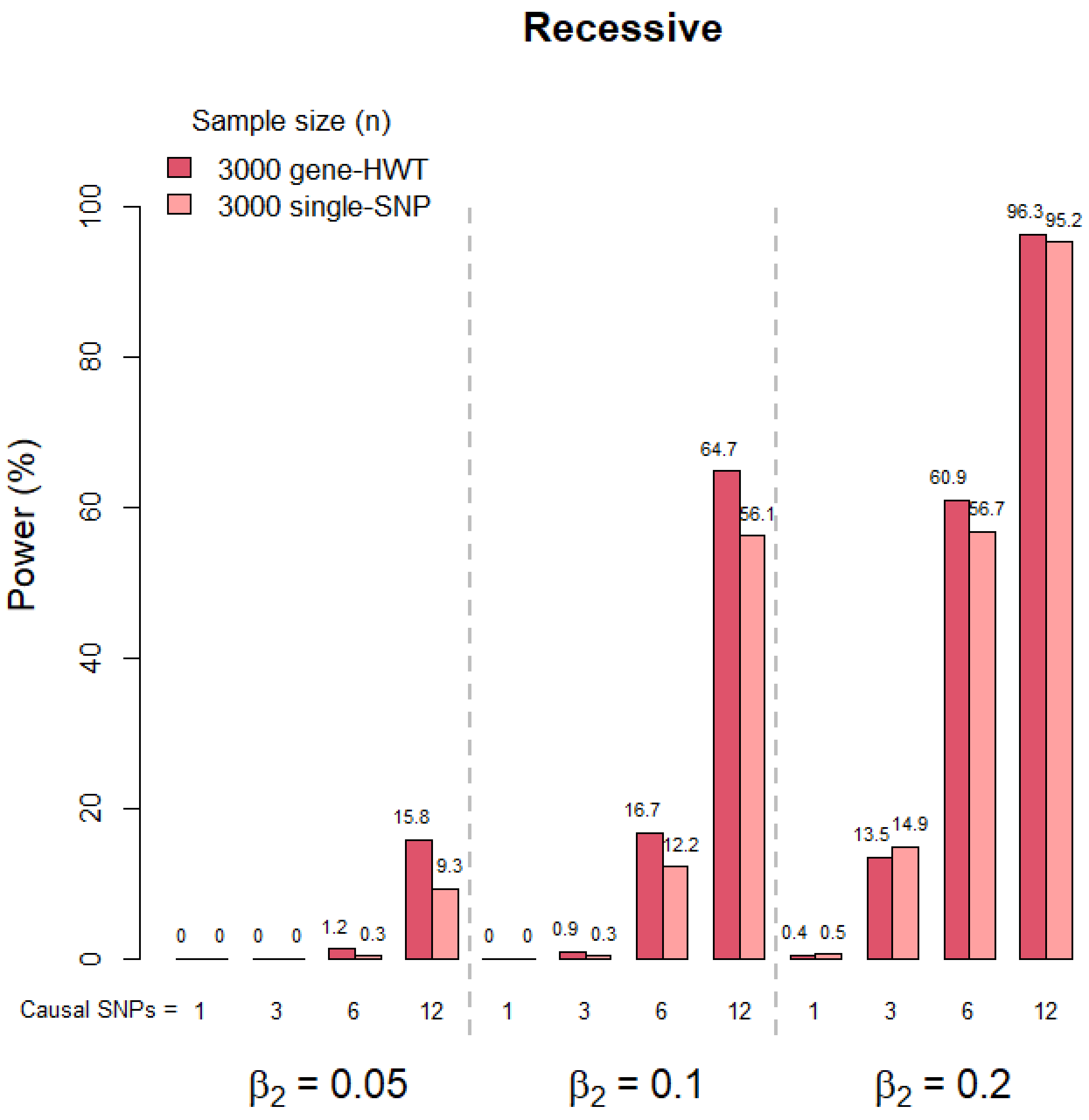

Supplementary Figure 2. Countineud.

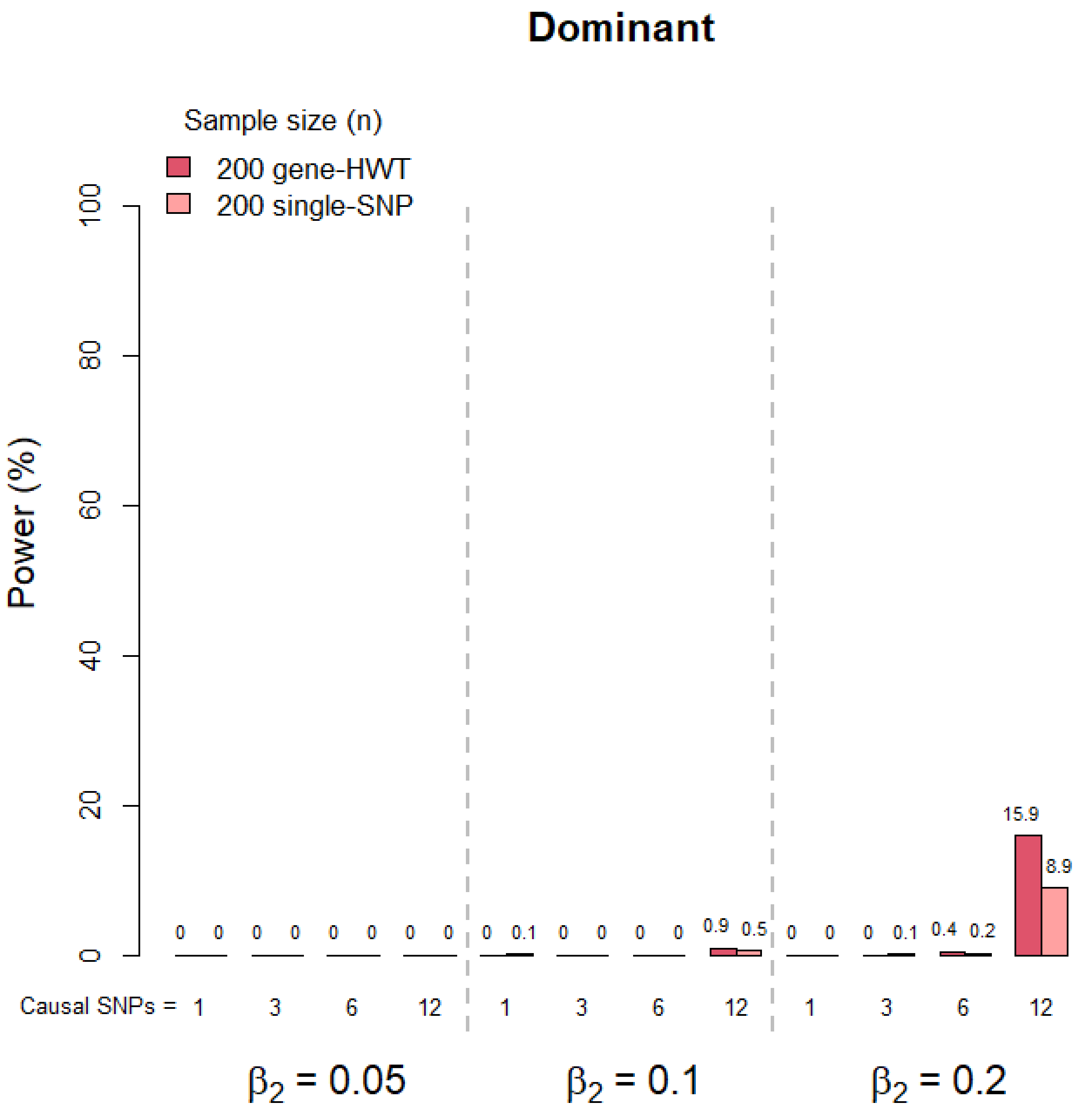

Supplementary Figure 2. Countineud.

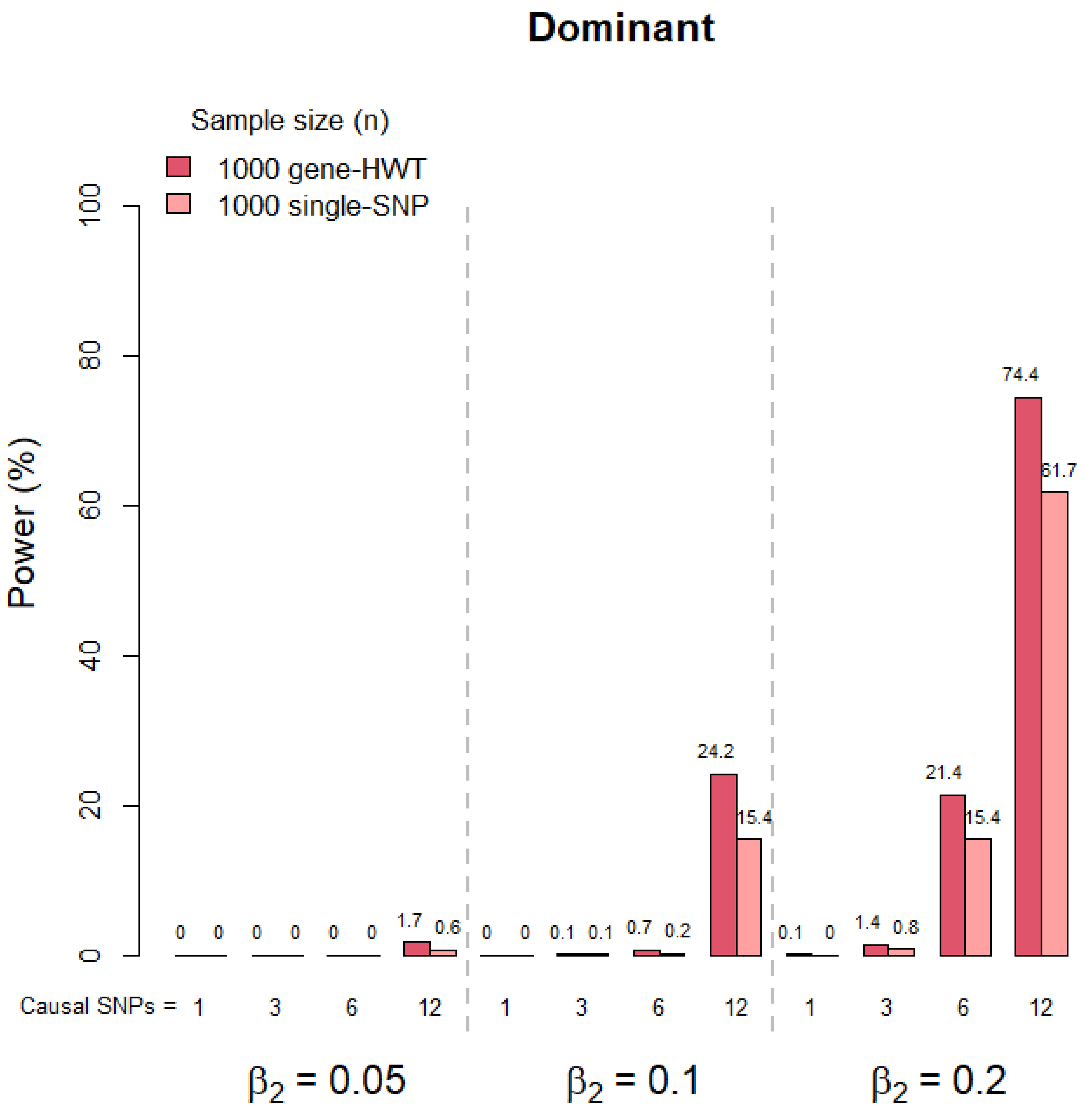

Supplementary Figure 2. Countineud.

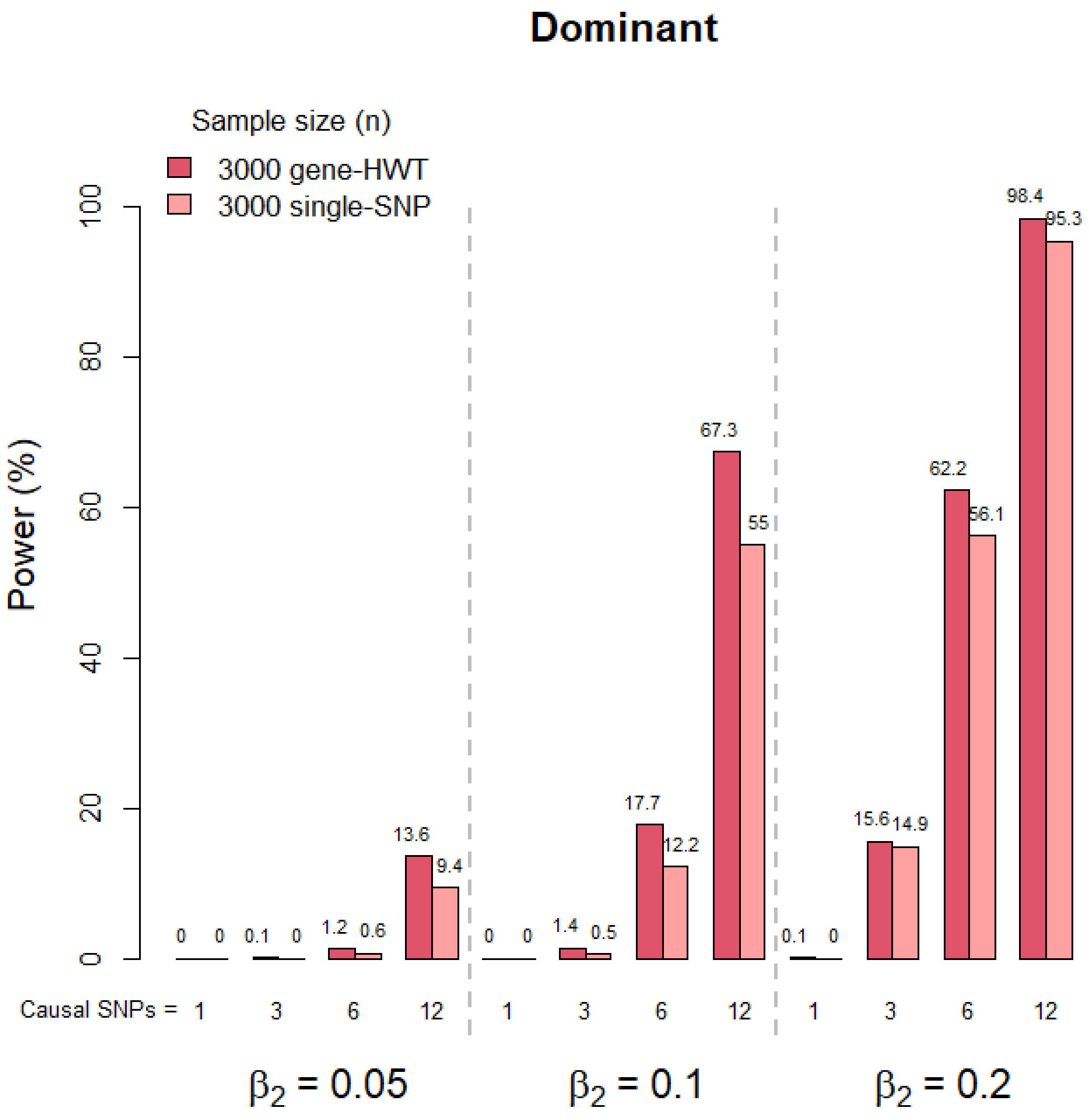
